# Deceased donor kidney function is determined by branch chained amino acid metabolism during ex vivo normothermic perfusion

**DOI:** 10.1101/2023.11.15.23298543

**Authors:** Armin Ahmadi, Jacquelyn Yu, Jennifer E. Loza, Brian C. Howard, Ivonne Palma, Peter A. Than, Naeem Makarm G Goussous, Junichiro Sageshima, Baback Roshanravan, Richard V. Perez

**Affiliations:** Department of Medicine, Division of Nephrology, University of California, Davis, CA, USA; Department of Surgery, Division of Transplant, University of California Davis Health, Sacramento, California, USA

**Keywords:** Normothermic perfusion, amino acid metabolism, transplantation

## Abstract

Current kidney perfusion protocols are not optimized for addressing the ex vivo physiological and metabolic needs of the kidney. Ex vivo normothermic perfusion (EVNP) may be utilized to distinguish high-risk kidneys to determine suitability for transplantation. We assessed the association of tissue metabolic changes with changes in kidney injury biomarkers and functional parameters in eight deceased donor kidneys deemed unsuitable for transplantation during a 12- hour ex vivo normothermic perfusion (EVNP). The kidneys were grouped into good and poor performers based on blood flow and urine output. The mean age of the deceased kidney donors was 43±16 years with an average cold ischemia time of 37±12 hours. Urine output and creatinine clearance progressively increased and peaked at 6 hours post-perfusion among good performers. Poor performers had 71 ng/ml greater (95% CI 1.5, 140) urinary neutrophil gelatinase-associated lipocalin (NGAL) at 6 hours compared to good performers corresponding to peak functional differences. Organ performance was distinguished by tissue metabolic differences in branch-chained amino acid (BCAA) metabolism. Tissue BCAA levels negatively correlated with urine output among all kidneys at 6 hours. Tissue lipid profiling showed poor performers were highlighted by the accumulation of membrane structure components including glycerolipids and sphingolipids at early perfusion time points. Overall, we showed that 6 hours is needed for kidney functional recovery during ENVP and that BCAA metabolism may be a major determinant of organ function and resilience.

## Introduction

The most significant obstacle in kidney transplantation is the shortage of available donor organs. The number of patients waiting continues to increase, from nearly 19,000 in 2000 to over 100,000 in 2019 ^1,2^. To address this, the deceased donor pool has expanded to include higher-risk organs such as those with acute kidney injury, donation after circulatory death, and older donors with defined comorbidities ^3^. By expanding the donor pool, the frequency of kidney discards has also increased despite evidence to suggest that kidneys with similar characteristics have been successfully transplanted with good outcomes. The 2021 Organ Procurement and Transplantation Network (OPTN) report states that over 18,000 deceased donor kidney transplants were performed with approximately 3,600 kidneys recovered being discarded ^4^. It has been postulated that a substantial number of discarded kidneys would be transplanted if better modalities to predict successful outcomes existed ^5^. Current preservation modalities are limited in their ability to accurately predict which of these high-risk discarded organs could have been transplanted with ultimate good function. As the need for organs for transplantation increases, the need for improved assessment and preservation strategies is more evident.

Current preservation techniques are based on hypothermic conditions to decrease metabolic activity and minimize cellular injury. However, there is no clear-cut way to pre-operatively assess the ultimate viability and function of the hypothermic kidney in a relatively inert metabolic state prior to transplantation. Ex vivo normothermic machine perfusion (EVNP) is another organ preservation system that poses several advantages such as the ability to monitor kidney blood flow, function and metabolism prior to transplantation and support repair of reversible injury ^6–8^. By re-establishing normal metabolism using a blood-based perfusate, EVNP can replenish energy stores with increased adenosine triphosphate (ATP) levels and improve renal allograft function after ischemia-induced injury ^9–12^. It can also potentially reduce the injury caused by cold ischemia. Several studies have used EVNP clinically and found improved allograft function and a decrease in frequency of delayed graft function ^6, 9,13–16^. These initial clinical reports have not yet determined the optimal conditions for EVNP including perfusate composition and duration of perfusion.

All deceased donor transplantations require a period of ischemia and hypoxemia during hypothermic preservation, contributing to some degree of Acute Kidney Injury (AKI). AKI during deceased donor preservation and transplantation is purported to involve mitochondrial dysfunction characterized by increased reactive oxygen species (ROS), and impaired fatty acid oxidation (FAO) and reduced ATP generation^17,18^. We performed untargeted metabolomics and lipidomics of from serial renal cortical tissue biopsies to better characterize metabolic derangements occurring in the setting of AKI during deceased donor kidney preservation. Our goals are twofold— to perfuse kidneys for an extended period to gain insight into optimal perfusion time and to characterize the link between kidney function and metabolic competence during a 12-hour ex vivo normothermic perfusion period.

## Methods

### Deceased donor kidneys

Eight human kidneys from eight deceased donors deemed unsuitable for transplantation were included in this study and transported on ice to the Transplant Surgery research laboratory at the University of California, Davis Health. The kidneys were preserved using 12 hours of EVNP during which hemodynamics and markers of perfusion, function, and injury were monitored. Kidney cortex tissue samples were also obtained throughout the perfusion period and sent for metabolomics and lipidomics analysis (West Coast Metabolomics Center, Davis, CA). This study was approved by the University of California, Davis Institutional Review Board, and was considered exempt from informed consent. This study was also approved by our institution’s Human Anatomical Specimen and Tissue Oversight Committee.

### EVNP system

All kidneys were placed on an EVNP system using a pediatric cardiopulmonary bypass circuit with an oxygen reservoir (Terumo CAPIOX RX05) and centrifugal pump (Maquet Rotaflow) as previously described ^19^. Briefly, normothermia was maintained at 37°C with a heat exchanger (Medtronic BIOCAL 370). The perfusate was pumped into the kidney via the cannulated renal artery and venous output was passively collected in a custom stainless steel kidney chamber and returned to the circuit. The kidneys were dissected free of perirenal fat tissue to allow for clear viewing during perfusion. The renal artery was dissected out and cannulated with a stainless-steel cannula (3 to 6 mm, Waters Medical System, Rochester, MN), and the renal vein was left to drain passively into the reservoir. Flow and pressure monitors were used to maintain mean arterial pressure at 70 to 80 mmHg. The ureters were cannulated using a five-French feeding tube (Covidien, Minneapolis, MN) for urine collection. The perfusate was continuously oxygenated with oxygen/carbon dioxide gas (95%/5%, 2 L/min) resulting in average pO_2_ levels of 580 mmHg over the perfusion period.

### Composition of EVNP perfusate

Kidneys were perfused with a blood-based perfusate consisting of leukocyte and platelet depleted, washed packed red blood cell (PRBC) units blood type O positive, which were acquired from our internal hospital blood bank and diluted in a 1:1 ratio with Plasmalyte-A (Baxter Medical, Deerfield, IL).

The perfusate was supplemented with heparin (2000 international units, NOVAPLUS), exogenous anhydrous creatinine (0.06 g, MP Biomedicals, Burlingame, CA), 20 ml of parenteral nutrition (Baxter CLINIMIX E 2.75/10) which was infused with 100 units of regular insulin (Humulin R), 5 ml of multivitamins (Baxter Medical), and 26 ml of 8.4% Bicarbonate (Millipore Sigma, St. Louis, MO). The parenteral nutrition was continued at the rate of 20 ml/hour until the end of the perfusion period.

The circuit was primed by re-circulation of the prepared PRBC perfusate oxygenated with 95% O_2_ /5% CO_2_ gas mixture for one to two minutes. The kidneys were then removed from the ice and placed on the stainless-steel chamber, connecting the arterial line to the cannula ensuring no air. To prevent desiccation, the kidney was covered with plastic wrap and the surface irrigated with Plasmalyte-A. Urine loss was replaced at a 1:1 ratio with Plasmalyte-A.

### Kidney assessment

Hemodynamic parameters were recorded every 30 minutes and intrarenal resistance (RR) was calculated as pressure (mmHg)/ flow (ml/min). Urine was collected every 30 minutes and centrifuged at 1500 rpm for five minutes (Sorvall RT7 Plus Centrifuge) and stored in a -20°C freezer for further analysis. Urine samples were later analyzed for sodium using a Critical Care Xpress Machine (Nova Biomedical, Waltham, MA) and for creatinine using a creatinine parameter assay kit (R&D systems, Minneapolis, MN) for calculation of creatinine clearance and fractional excretion of sodium (FENa) as reported previously ^20^. Urinary neutrophil gelatinase-associated lipocalin (NGAL) was measured with a point-of-care fluorescence immunoassay system (Bio Site Triage MeterPro Alere, Waltham, MA).

Perfusate samples were collected every 30 minutes from a port in the arterial line and directly from the venous return. The samples were analyzed by a Critical Care Xpress machine for pH, pCO_2_, and electrolytes, and handheld point-of-care devices (Stat sensors, Nova Biomedical) for creatinine, lactate, and glucose levels. Based on arterial pH values, an additional 8.4% bicarbonate was added to the reservoir to correct for low pH and achieve physiological pH of 7.3-7.4.

The kidneys were grouped into good and poor performers based on global appearance and functional parameters including renal blood flow and urine output similar to previous studies ^13,14,20^. Briefly, kidneys that on initiation of EVNP perfused uniformly and quickly with a pink appearance throughout, attained a renal blood flow over 150 ml/min, and produced urine greater than 20 ml over 30 minutes were designated as the “good performers”. Kidneys that had a slower reperfusion with patchy coloration and areas of poor perfusion, with blood flow less than 150 ml/min and urine output less than 20 ml/30 minutes were designated “poor performers”.

### Metabolomics Profile

Renal cortical core tissue needle biopsies (16g Tru Core II Biopsy Instrument, Argon Medical Devices, Athens, TX) were taken prior to and at various time points over 12 hours of EVNP, snap frozen in liquid nitrogen, and stored in a -80°C freezer. After study completion, samples were submitted to the West Coast Metabolomics Center at the University of California Davis. Samples were analyzed with validated chromatography-mass spectrometry (GC-TOF) for an untargeted primary metabolites analysis. A total of 124 unique metabolites were detected using this method.

### Lipidomics profile

The lipidomics workflow involved sample extraction in MTBE with addition of internal standards based on the “Maytash” method ^21^, followed by ultra-high-pressure liquid chromatography (UHPLC) on a Waters CSH column, interfaced to a quadruple time-of-flight (QTOF) mass spectrometer (high resolution, accurate mass), with a 15-minute total run time. Data were collected in both positive and negative ion mode and analyzed using MassHunter (Agilent). A total of 311 unique lipid species were detected.

### Statistical Analysis

A retrospective analysis was done to determine group assignment of the discarded kidneys into poor and good performers. We used ANOVA adjusted for multiple hypothesis testing (Bonferroni) to determine significant differences in hemodynamics and metabolites over time. To assess changes in tissue metabolism, a paired t-test was performed to determine significantly altered metabolites between good and poor performers at various time points over 12 hours. Spearman correlation coefficient was used to assess the correlation of tissue BCAAs with urine output. Metaboanalyst 5.0 was used for enrichment analysis using significantly altered metabolites between good and poor performers. All tests were two-sided and p-values < 0.05 were concluded statistically significant. Analyses were performed with SAS software, version 9.4 (SAS institute, Cary, NC). GraphPad Prism 9.0 (GraphPad Software, Inc., San Diego, California) was used to generate the graphs.

## Results

### Donor characteristics

Individual donor characteristics, reasons for declined clinical use, and their retrieval conditions prior to perfusion are summarized in **Table 1**. The age of the 8 deceased kidney donors ranged from 17 to 65 years with a mean of 43 ± 16 years. Two out of the total 8 kidneys were from female donors. The average body weight of the donors was 94 ± 28 kg. None of the donors were diabetics while 5 out of 8 donors had hypertension. The average cold ischemia (CIT) was 37 ± 12 hours (range 25- 56 hours).

**Table 1.** Individual kidney donor characteristics and reasons for discard.

|  | **Age range (years)** | **Sex** | **Weight (kg)** | **Donor type** | **CIT (hours)** | **Hypertension** | **Cause of death** |
| --- | --- | --- | --- | --- | --- | --- | --- |
| **Kidney 26** | 61-65 | M | 93 | DBD | 39 | Yes | Head trauma/Blunt injury |
| **Kidney 27** | 51-55 | F | 56 | DBD | 28 | Yes | CVA/Stroke/IH |
| **Kidney 33** | 41-45 | M | 91.3 | DBD | 38 | Yes | CVA/Stroke/IH |
| **Kidney 35** | 41-45 | M | 106.5 | DBD | 56 | No | Head trauma/stroke |
| **Kidney 36** | 51-55 | M | 94.2 | DBD | 54 | Yes | CVA/stroke |
| **Kidney 37** | 21-25 | F | 68 | DBD | 33 | No | Head trauma/Gun shot |
| **Kidney 38** | 46-50 | M | 151 | DBD | 25 | Yes | CVA/Stroke |
| **Kidney 39** | 16-20 | M | 90.7 | DCD | 25 | No | Anoxia |

DCD, donation after circulatory death; DBD, donation after brain death; CIT, cold ischemia time.

**Table 2** summarizes the characteristics of poor performers versus good performers assigned based on kidney perfusion parameters and urine output. Both groups had comparable sex composition and cold ischemia time (**Table 2**). On average, poor performers were younger, weighed less, had less hypertension and lower Kidney Donor Profile Index (KDPI) scores (**Table 2**).

**Table 2.**
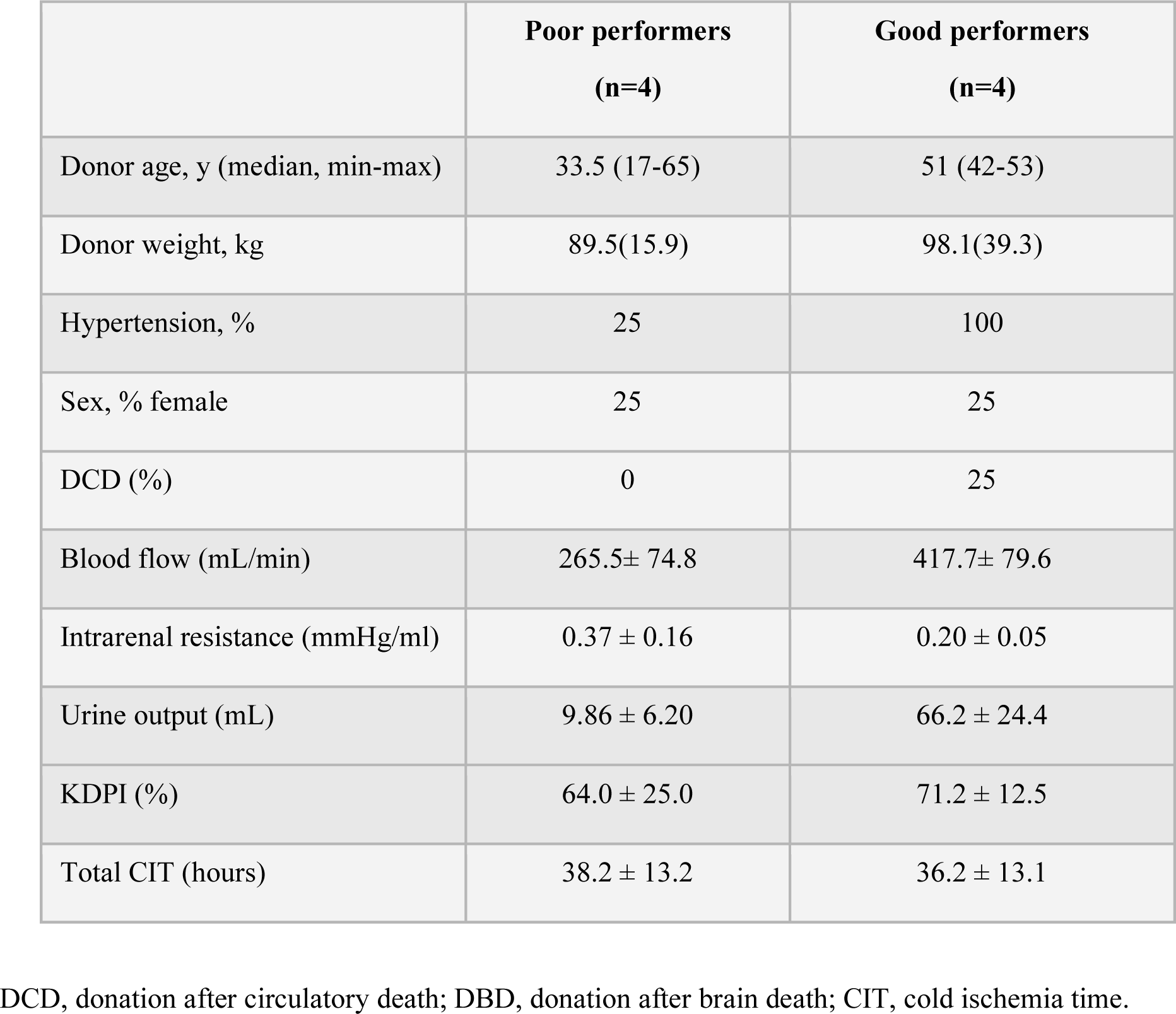
Grouped donor characteristics, hemodynamics, and perfusion characteristics.

|  | **Poor performers**<br>**(n=4)** | **Good performers**<br>**(n=4)** |
| --- | --- | --- |
| Donor age, y (median, min-max) | 33.5 (17-65) | 51 (42-53) |
| Donor weight, kg | 89.5(15.9) | 98.1(39.3) |
| Hypertension, % | 25 | 100 |
| Sex, % female | 25 | 25 |
| DCD (%) | 0 | 25 |
| Blood flow (mL/min) | 265.5± 74.8 | 417.7± 79.6 |
| Intrarenal resistance (mmHg/ml) | 0.37 ± 0.16 | 0.20 ± 0.05 |
| Urine output (mL) | 9.86 ± 6.20 | 66.2 ± 24.4 |
| KDPI (%) | 64.0 ± 25.0 | 71.2 ± 12.5 |
| Total CIT (hours) | 38.2 ± 13.2 | 36.2 ± 13.1 |

DCD, donation after circulatory death; DBD, donation after brain death; CIT, cold ischemia time.

### Hemodynamic and functional parameters

A summary of grouped hemodynamic and functional parameters is shown in (**Figure 1**). On average, poor performing kidneys had lower kidney blood flow and urine output coinciding with greater values for urinary NGAL as compared to good performing kidneys. In addition, we tracked the changes in additional parameters including creatinine clearance over the 12-hour perfusion period (**Figure 1A-F**). Poor performers had lower blood flow with a mean AUC of 3201 ml/min*hour (95% CI 2820 to 3581) compared to good performers with an AUC 5064 ml/min*hour (95% CI 4373 to 5756) **(Figure 1A**). In addition, poor performers had higher interrenal resistance with a mean AUC of 4.3 mmHg/ml*hour (95% CI 3.3 to 5.2) compared to a mean AUC of 2.3 mmHg/ml*hour (95% CI 2.0 to 2.6) among good performers **(Figure 1B)**. Urine output was higher among good performers at with an AUC (95% CI) of 820 ml*hour (95% CI 672 to 969) compared to an AUC of 122 ml*hour (95% CI 87 to 157) among poor performers **(Figure 1C**). Urinary NGAL levels were also elevated among poor performers with an AUC of 984 (95% CI 827 to 1100) compared with an AUC of 284 ng/ml*hour (95% CI 216 to 352) among good performers **(Figure 1D)**. Creatinine clearance was higher throughout the perfusion period among good performers with an AUC of 229 (95% CI 152 to 306) compared to an AUC of 18 ml*hour (95% CI 14 to 20) among poor performers (**Figure 1E**).

**Figure 1.**
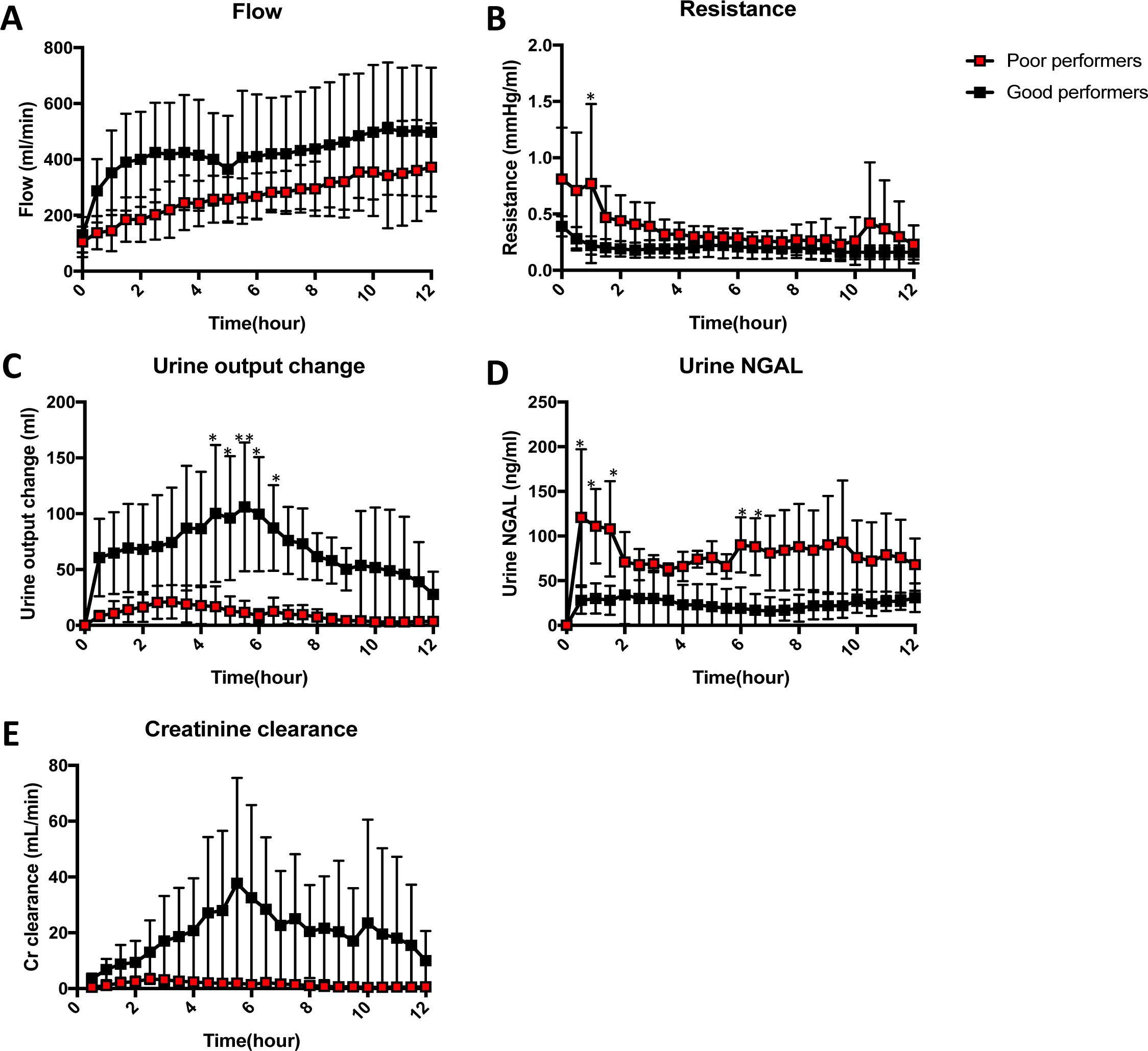
The hemodynamics among poor (n=4) and good performers (n=4) over the 12-hour perfusion period. A) urinary flow, B) intrarenal resistance, C) change in urine output, D) concentration of urinary NGAL, and E) creatinine clearance. Data points represent mean and error bars represent SD. “***” = p-value<0.001, “**” = p-value<0.01, “*” = p-value<0.05.

### Tissue metabolic profiling

During the initial 5 hours of perfusion the metabolic profiles between the two groups were similar. Thereafter, a total of 18 (14%) metabolites were significantly altered between the two groups at 6 hours and remained altered for 3 hours before converging at the end of the 12-hour perfusion period (**Figure 2**, **Table 3**). These changes predominantly involved elevations in amino acids (8/18 altered metabolites) in good performing kidneys relative to poor performing kidneys, including all branched chain amino acids (BCAA), valine, leucine, and isoleucine (**Table 3**). In addition to amino acids, tissue urea (end-product of amino acid catabolism) levels were also significantly higher among poor performers with a fold change of 1.44 (p-value of <0.01) (**Table 3**). The metabolites with the largest fold changes between good and poor performers included leucine, isoleucine, and histidine with fold changes of 2.2, 2.4, and 2.3 (*p*-value of <0.01 for all three) (**Table 3**). Enrichments analysis of the significantly altered metabolites at 6-hours revealed impaired amino acid metabolism (especially BCAA metabolism) as the dominant altered metabolic pathway among poor performers (**Supplemental Figure 1**).

**Figure 2.**
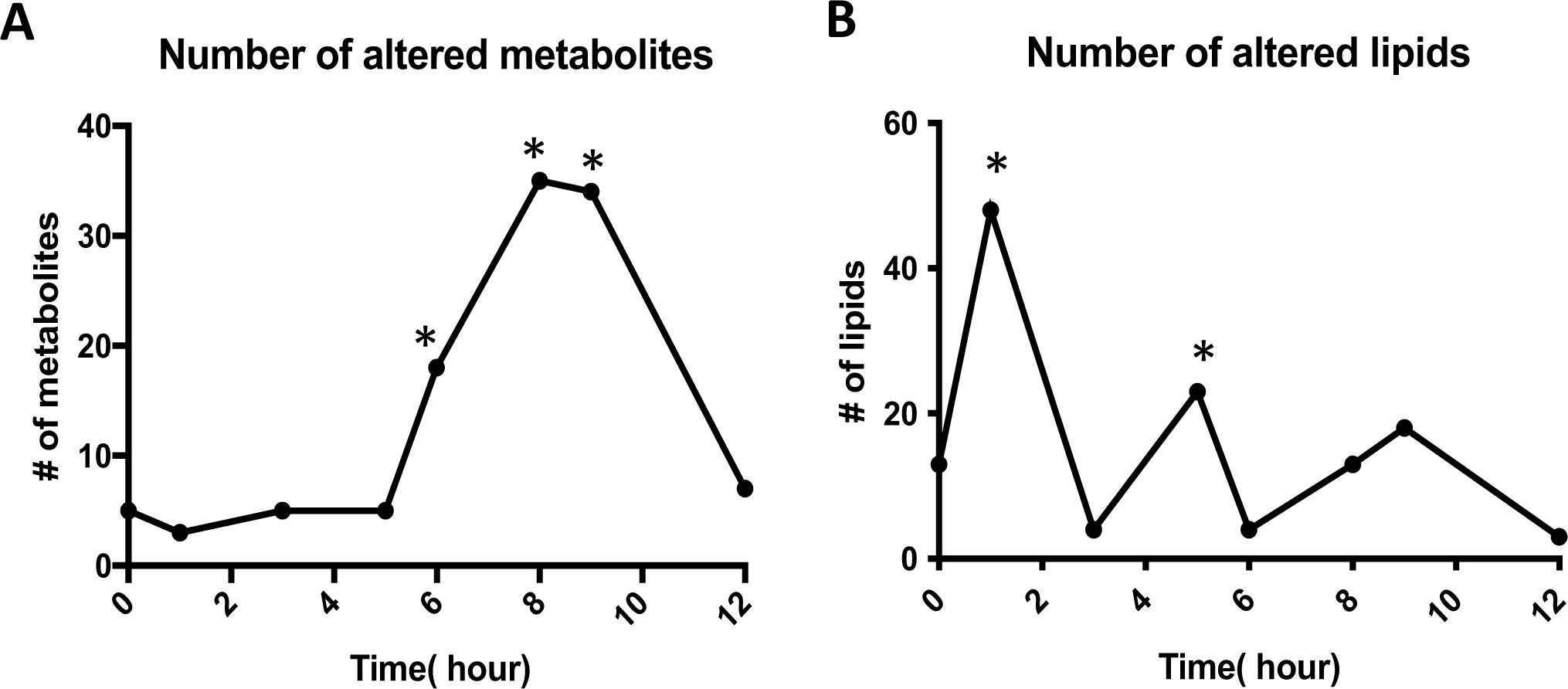
**Metabolic and lipid profile alterations compared to good performers. (**A) number of significantly altered (A) metabolites and (B) lipids. “*” represents significant alterations in at least 5% of the total detected metabolites/lipids.

**Table 3.**
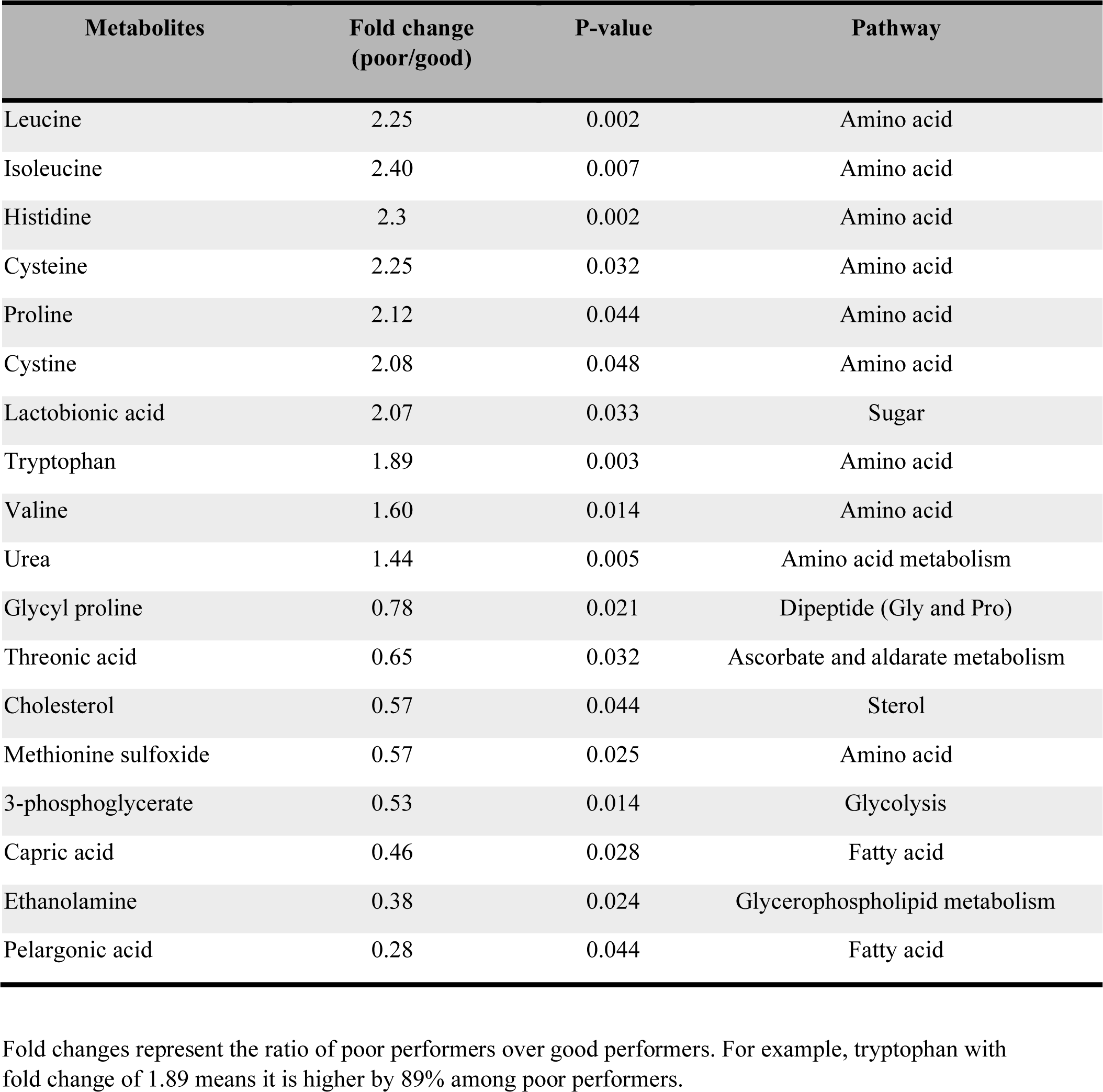
Differences in tissue metabolites at 6-hours during the perfusion period comparing poor (n=4) vs good performers (n=4).

| Metabolites | Fold change (poor/good) | P-value | Pathway |
| --- | --- | --- | --- |
| Leucine | 2.25 | 0.002 | Amino acid |
| Isoleucine | 2.40 | 0.007 | Amino acid |
| Histidine | 2.3 | 0.002 | Amino acid |
| Cysteine | 2.25 | 0.032 | Amino acid |
| Proline | 2.12 | 0.044 | Amino acid |
| Cystine | 2.08 | 0.048 | Amino acid |
| Lactobionic acid | 2.07 | 0.033 | Sugar |
| Tryptophan | 1.89 | 0.003 | Amino acid |
| Valine | 1.60 | 0.014 | Amino acid |
| Urea | 1.44 | 0.005 | Amino acid metabolism |
| Glycyl proline | 0.78 | 0.021 | Dipeptide (Gly and Pro) |
| Threonic acid | 0.65 | 0.032 | Ascorbate and aldarate metabolism |
| Cholesterol | 0.57 | 0.044 | Sterol |
| Methionine sulfoxide | 0.57 | 0.025 | Amino acid |
| 3-phosphoglycerate | 0.53 | 0.014 | Glycolysis |
| Capric acid | 0.46 | 0.028 | Fatty acid |
| Ethanolamine | 0.38 | 0.024 | Glycerophospholipid metabolism |
| Pelargonic acid | 0.28 | 0.044 | Fatty acid |

Fold changes represent the ratio of poor performers over good performers. For example, tryptophan with fold change of 1.89 means it is higher by 89% among poor performers.

### Temporal associations of tissue metabolites

After adjusting for multiple hypothesis testing, only temporal differences in BCAA were detected during the 12-hour period (**Figure 3, Supplemental Figure 2**). Poor performers had significantly higher levels of leucine and isoleucine starting at 6 hours and lasting until 9 hours post perfusion (**Figure 3A and B**). Similarly, valine also trended higher among poor performers at 6 (*p*-value of 0.10), 8, and 9 hours (**Figure 3C**) but did not reach statistical significance. All three BCAAs showed a significant negative correlation with urine output across all kidney donors at 6 hours. The strongest correlation belonged to leucine and isoleucine (r = -0.81 and *p*-value of 0.02 for both) followed by valine (r = -0.78, *p*-value of 0.03) (**Figure 4**).

**Figure 3.**
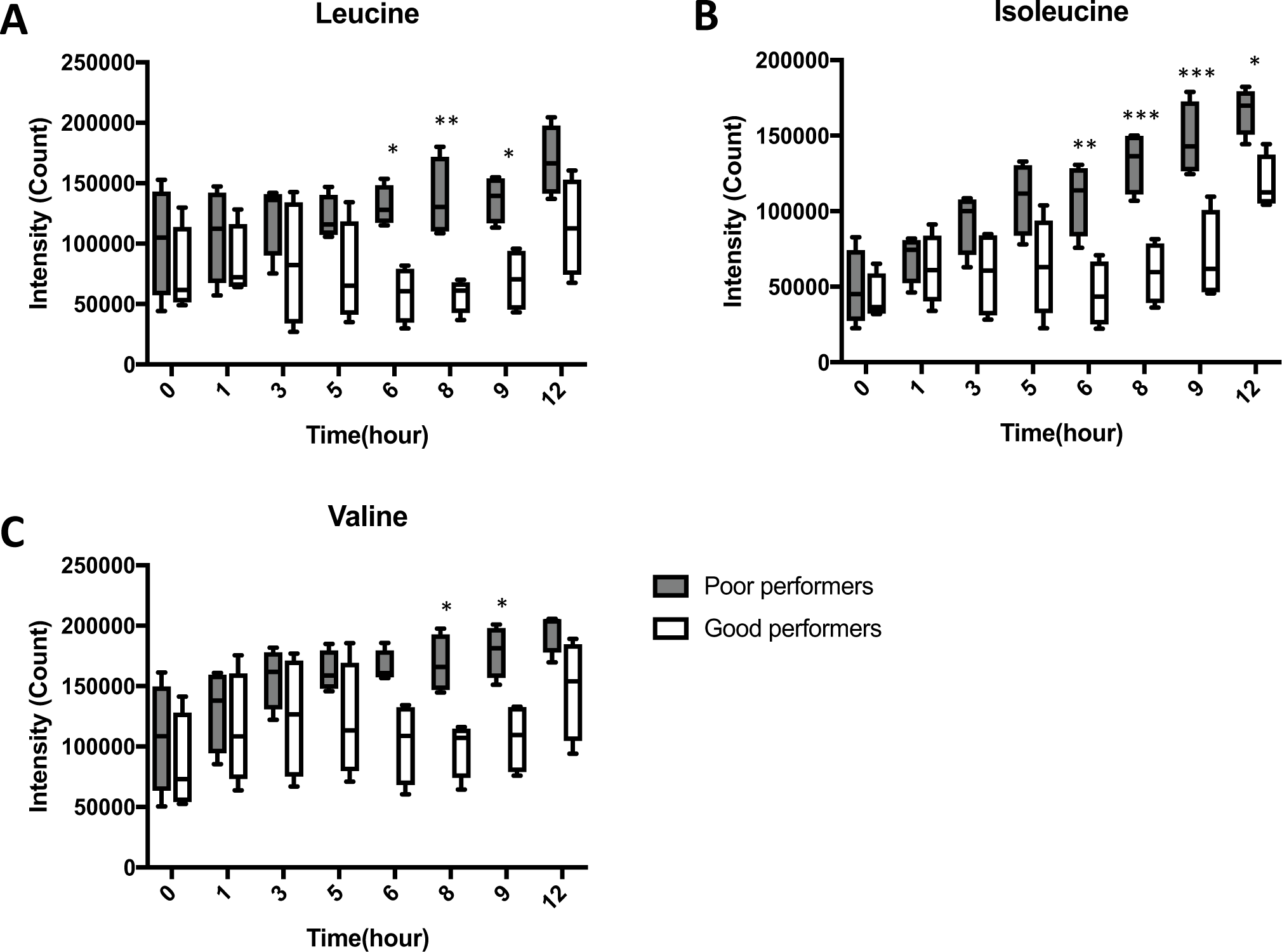
**Changes in tissue BCAA’s levels comparing poor (n=4) and good performers (n=4) over the 12-hour perfusion period**. A) Leucine, B) isoleucine, and C) valine are shown. The box plots represent median (min to max), and the error bars represent 95% CI. “***” = p-value<0.001, “**” = p-value<0.01, “*” = p-value<0.05.

**Figure 4.**
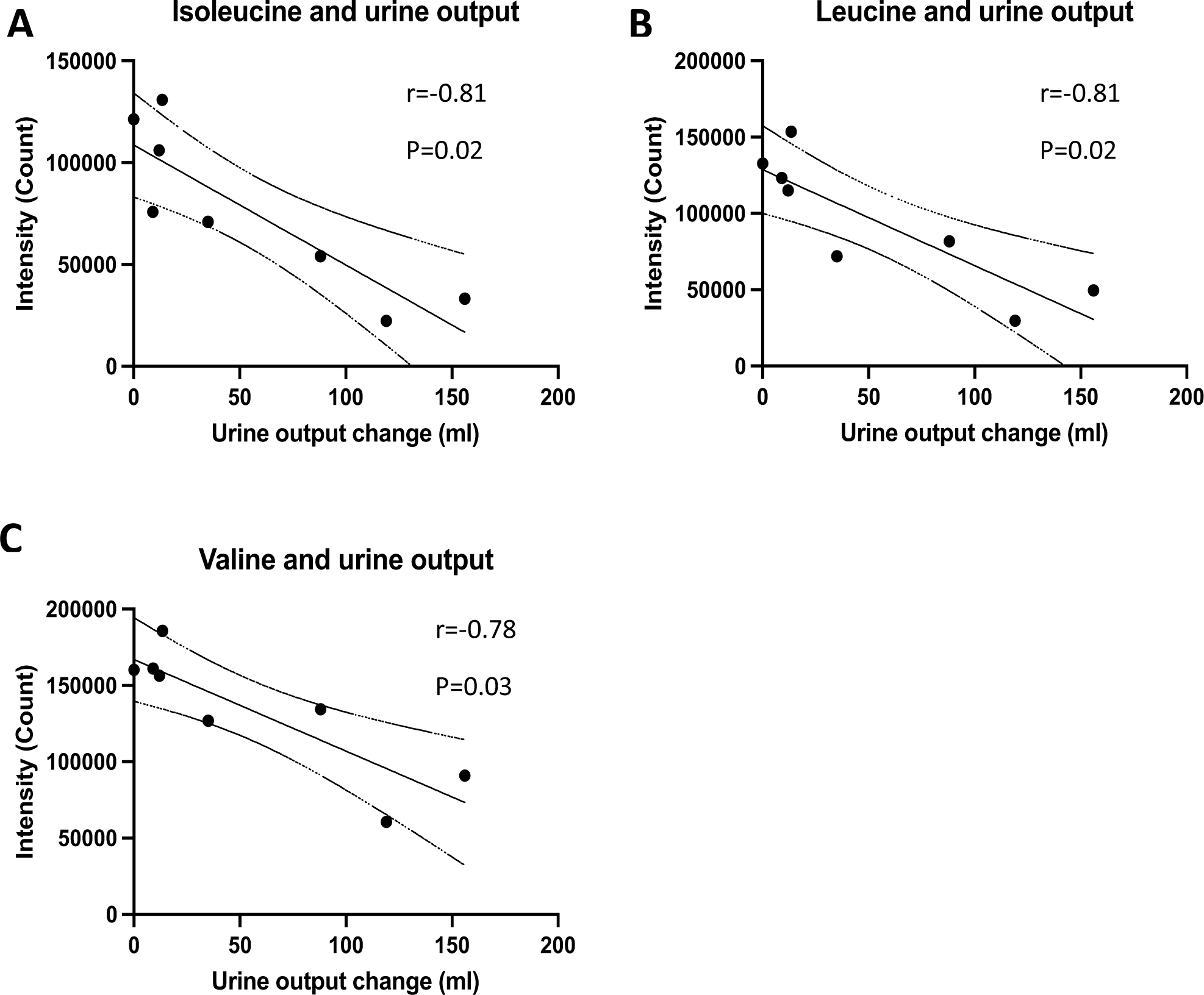
Cross-sectional correlation between BCAA’s and urine output at 6 hours post perfusion (n=8). The correlation between tissue A) Leucine, B) isoleucine, and C) valine are shown.

To assess if alterations in tissue metabolic profile were influenced by reduced excretion or reabsorption, we evaluated differences in altered tissue to urine metabolite ratios comparing good and poor performers at 6 hours. We found no overlap between the altered tissue and urine metabolites (**Supplemental Table 1**). We found that the urine to tissue ratio of BCAA’s were higher in good performers compared to the poor performers(**Supplemental Figure 3A-C**).

Restricting analysis among good performers to the period between 6-12 hours post perfusion corresponding to a decline from peak to trough levels of kidney function (**Figure 1C-F**) revealed 19 (15%) significantly altered metabolites. Within the subgroup of good performing kidneys declining kidney function from 6 to 12 hours coincided with elevations in cortical tissue amino acids including leucine and isoleucine. The metabolites with the highest fold changes comparing 12 hours to 6 hours included isoleucine (fold change of 2.6), proline, and ornithine (fold change of 2.14 for both) (**Supplemental Table 2**).

We evaluated other parameters of metabolic competence and viability including glucose consumption, glycolysis final products (pyruvate and lactate), TCA cycle intermediates, and 3- hydroxybutyrate as a marker for beta oxidation rate (**Figure 5**). We found a sharp increase in glucose accumulation over time with significantly higher glucose levels at 9- and 12-hours post perfusion among poor performers compared to good performers (**Figure 5A**). There were no significant differences in 3-hydroxybutyrate, pyruvate, and lactate at any timepoint during perfusion (**Figure 5B-D**). We also found no difference in any of the detected TCA cycle intermediates at any timepoint (**Supplemental Figure 4**).

**Figure 5.**
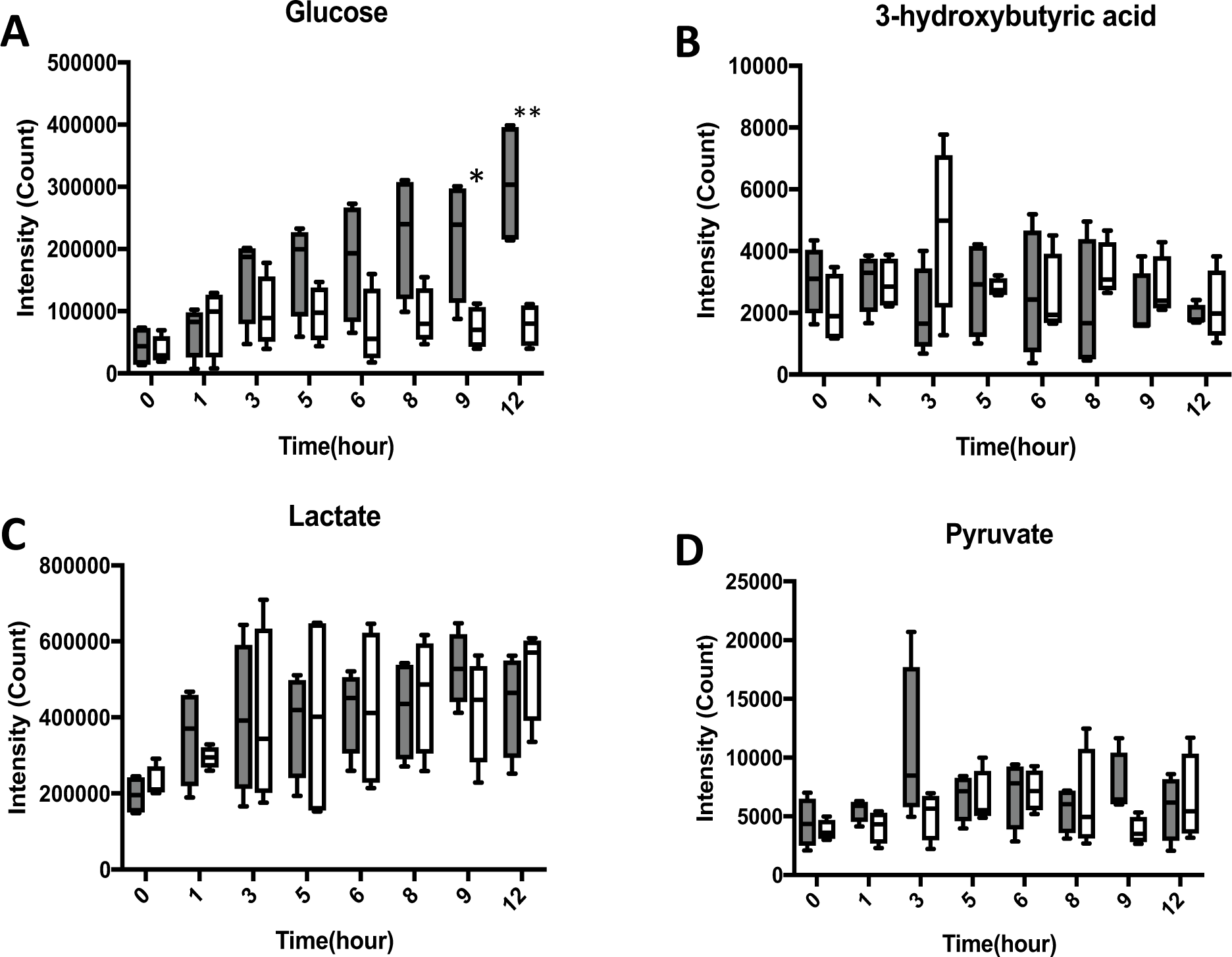
**Changes in tissue glucose, glycolysis products, and 3- hydroxybutyrate levels over the 12-hour perfusion period in both groups**. Grey bars represent poor performers and white bars represent good performers.

### Changes in tissue metabolome and temporal associations with kidney function

During the first 5 hours, less than 5% of detected metabolites significantly differed between good and poor performers (**Figure 2**). Substantial differences in metabolites were first noted at 6 hours, increasing to a peak differential metabolic profile by 9 hours post perfusion (**Figure 2A**). By 12 hours, these changes substantially attenuated (**Figure 2A**). Detectable differences in urine output and urinary NGAL between good and poor performers peaked at 6 hours coinciding with the onset of detectable differences in tissue metabolites (**Figure 1C and D**). Similar temporal trends were observed for differences in creatinine clearance and oxygen consumption (**Figure 1E and F**).

### Tissue lipid profiling

#### Changes in tissue lipid profile

In contrast to the metabolic profile, the peak number of altered lipid species distinguishing good and poor performers was observed at 1 hour post perfusion with 48 (19%) significantly altered lipids (**Figure 2A**). All 48 altered lipids were significantly elevated compared to good performers (**Figure 6 and Supplemental Table 3**), and they were predominantly composed of key plasma membrane/structure components including glycerolipids, sphingolipids, and fatty acyls (**Figure 6**). The largest fold changes at 1 hour belonged to triglycerides: TG (46:3), TG (60:2), and TG (58:3) (**Supplemental Table 3**).

**Figure 6.**
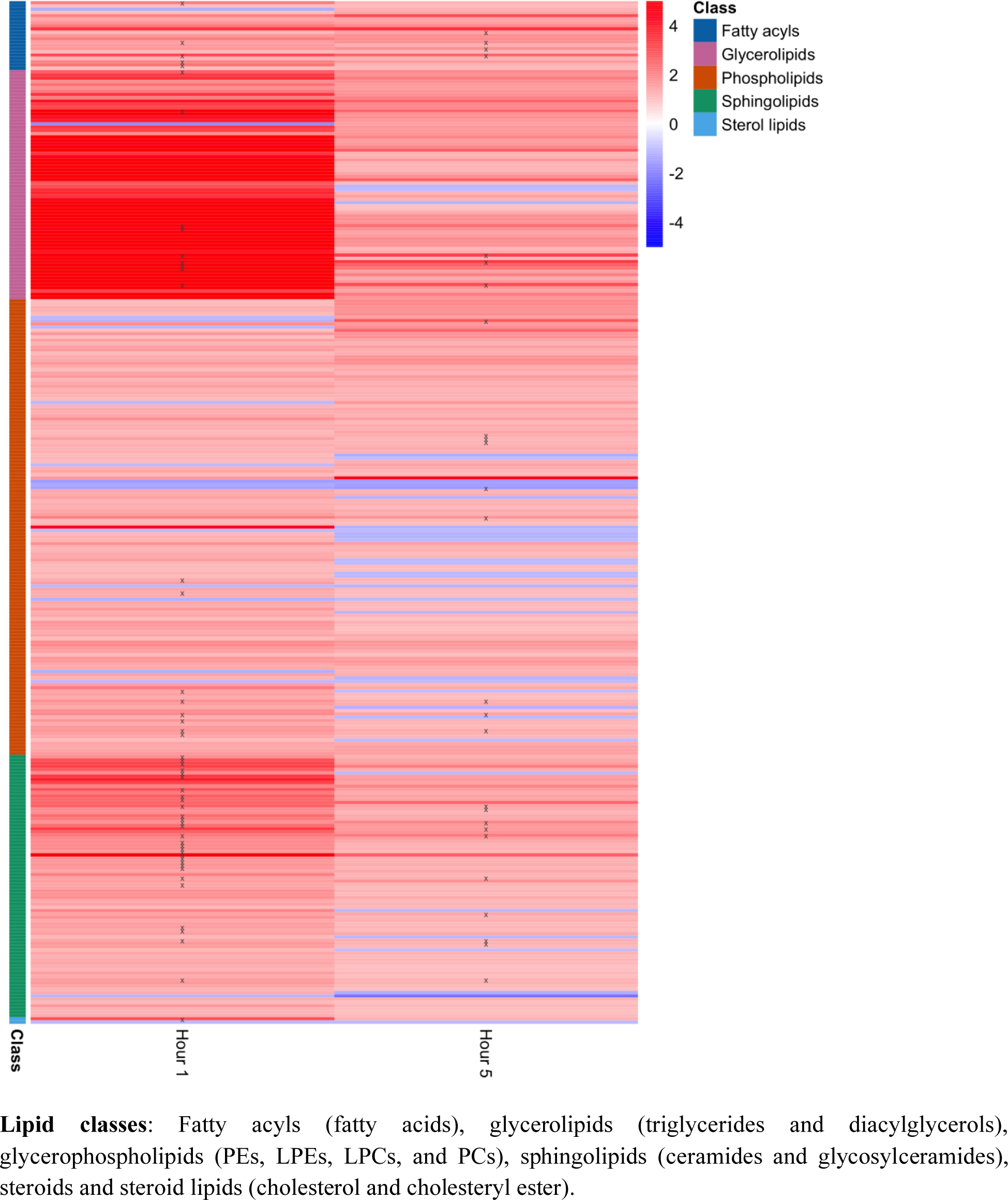
**Compositional changes within functional lipid classes at 1 hour and 5 hours post perfusion**. The colors of heatmap are based on fold changes obtained from the ratio of poor/good functioning groups. **P < 0.001, *P < 0.05.

The magnitude of difference in lipid profile distinguishing poor from good performing kidneys attenuated over time. At 5 hours, only 23 (9%) of the total detected lipid species were significantly different between the two groups, albeit at a lower magnitude compared to the first hour (**Figure 2A, Supplemental Table 4**). These lipids were primarily plasma membrane components including fatty acyls and sphingolipids (**Figure 6**). Similar to 1-hour changes, the lipid species with the largest fold changes at 5 hours involved triglycerides (**Supplemental Table 4**).

## Discussion

Our study applies an ex vivo normothermic perfusion (EVNP) system to assess hemodynamic, functional, and metabolic changes during a 12-hour kidney perfusion period to determine the optimal preservation time and identify tissue metabolic changes relating to parameters of kidney function. We demonstrate that the optimal perfusion duration is 6 hours with serial tissue metabolic profiling distinguishing poor performing kidneys at 6 hours by a pattern of impaired amino acid metabolism compared to good performing kidneys. In cross-sectional analysis tissue, BCAA levels negatively correlated with urine output among all kidneys at 6 hours. Tissue BCAA levels increased over time in poor performers compared with good performers. Poor performers were also distinguished by tissue glucose accumulation despite having comparable tissue TCA cycle intermediates, glycolysis end-products, and beta oxidation by-products. Lipid profiling showed poor performers accumulated tissue plasma membrane components such as glycerolipids and sphingolipids during early post perfusion timepoints. Overall, these findings provide insight into active lipid, carbohydrate, and protein metabolism during EVNP and suggest a key role for impaired BCAA metabolism in kidney functional decline.

Previous clinical studies focusing on the demonstration of safety and feasibility of EVNP have been performed with relatively short perfusion times in the setting of deceased donor kidney transplantation. Clinical reports from the United Kingdom (Hosgood), Canada ^16^, the Netherlands ^15^, Germany (Minor) and India (Choudary) all limit the perfusion time to 1-3 hours. One rationale for a brief period of normothermic perfusion is that restoration of cellular ATP homeostasis in deceased donor kidneys is achieved after EVNP for only 1-2 hours^27^. Our results suggest that continued normothermic perfusion up to 6 hours may offer additional benefit to the organ since functional parameters, including urine output and creatinine clearance, peak at this time point among the good performing kidneys (**Figure 1A-1E**). In our study, perfusion of greater than 6 hours resulted in function deterioration consistent with other studies^22^. Indeed, other preservation approaches using various physiological perfusion conditions to determine an optimal perfusion time has been used by others ^24,25^. Weissenbacher’s EVNP platform used a PRBC-based perfusate mixed with albumin and urine recirculation, which demonstrated that a human kidney can be perfused for up to 48 hours. The study showed that perfusion periods of more than 6 hours result in a more metabolically stable state with initiation of tissue recovery and down-regulation of markers of ischemia reperfusion injury ^23^. Indeed, perfusate composition is a major factor that directly impacts the functional and viability parameters inevitably changing optimal perfusion time ^24,28^. Graft viability assessment from kidney biopsies during EVNP show that higher renal blood flow is positively correlated with genes involved in oxidative phosphorylation pathways and negatively correlated with inflammatory gene pathways at 0- and 2-hour post perfusion^29^. This is in keeping with our functional data, showing higher renal blood flow and urine output with significantly lower urinary NGAL levels at 6-hour post perfusion when comparing good and poor performers.

Our tissue metabolic profiling showed impaired BCAA metabolism was linked to the decline of kidney function and hemodynamics during ENVP. A major component of the high energetic demand of the kidney proximal tube involves active re-absorption of filtered glucose and amino acid from circulating blood ^24,25^. The active transport machinery in the kidneys is driven by Na^+^/K^+^-ATPase located on the basolateral membrane of the tubules generating an electrochemical gradient, allowing for direct and indirect transport of amino acids and glucose ^26^. Thus, mitochondrial health and activity is vital in facilitating glucose and amino acid reabsorption and metabolism by the kidney^17^. This process allows for uptake, catabolism, synthesis, and release of amino acids that are vital in maintaining the integrity and physiology of the kidney ^25,27^ . The primary source of energy for the proximal tubes are free fatty acids ^28^, however our ENVP system was set up to assess metabolic plasticity to use glucose and (BCAA) amino acids as catabolic substrates. First, functional differences among good and poor performers were highlighted by impaired amino acid metabolism, specifically BCAAs at 6 hours post perfusion where functional differences were the largest between the two groups (**Figure 2**, **Table 3, Supplemental Figure 1**). Second, disturbances in BCAA metabolism were also shown in temporal comparison of tissue metabolic profile among good performers comparing 12-hour and 6-hour post perfusion (**Supplemental Table 2**). Third, our cross-sectional assessment showed a significant negative correlation between tissue BCAA levels with our primary functional parameter urine output at 6- hour post perfusion among all 8 kidneys (**Figure 4**). This suggests that mitochondrial dysfunction leads to limited metabolic plasticity of the kidney to metabolize amino acids. Our findings compliment those of a recent study of 190 kidney transplants post-hypothermic machine perfusion identified amino acid transport as one of the top altered metabolic pathways associated with death-censored delayed graft failure ^29^. Overall, our serial metabolic profiling suggests that BCAA oxidation is a crucial metabolic adaptation in maintaining kidney function during ENVP. Future studies with larger sample sizes are needed to study the link between BCAA metabolism and kidney viability and function during an ENVP.

Metabolic fitness is critical to tissue viability. This was assessed by measuring changes in glucose, glycolysis end-products, and TCA cycle intermediates. Glucose and amino acids are actively reabsorbed through an energy demanding process in the kidney ^30^. Glucose metabolism imposes an energetic burden on the kidneys, especially the proximal tubes during EVNP. We found marked and consistent accumulation of tissue glucose levels among poor performers while its levels were stable among good performers (**Figure 5A**). Glucose levels were significantly higher among poor performers at 9-hour and 12-hour post perfusion compared to good performers, signaling reduced glucose consumption among poor performers (**Figure 5A**). Despite higher tissue glucose levels, there was no difference between the two groups in the level of the important glycolysis end-products lactate and pyruvate, suggesting similar glycolytic activity (**Figure 5**). Of the four detected TCA cycle intermediates (succinate, fumarate, malate, and citrate) only malate and fumarate showed a declining pattern among poor performers compared to good performers (**Supplemental Figure 4**). Overall, our findings suggest metabolic plasticity characterized by both glucose and BCAA substrates is central to preserving kidney function during 12-hour ENVP.

Lipid metabolism during EVNP exhibited a different temporal profile when compared to amino acid metabolism. In contrast to changes in amino acid metabolism between groups which became evident later in the perfusion period, we found the largest lipid profile differences at 1-hour post perfusion comparing good and poor performers (**Figure 2**). These lipids involved significant accumulation of membrane structure components such as glycerolipids (triglycerides and diacylglycerols), sphingolipids (ceramides), and fatty acyls (fatty acids) among poor performers ^31,32^ (**Figure 6**). These differences persisted at 5 hours post perfusion albeit attenuated (**Figure 6, Supplemental Table 4**). This suggests that poor performers are associated with substantial tissue damage resulting in membrane breakdown and cell remodeling at early timepoints during ENVP. This is in line with other studies showing that hyperlipidemia including hypertriglyceridemia is strongly linked with development and progression of long-term allograft dysfunction ^33^. A previous study has also shown high fatty acid accumulation during hypothermic machine perfusion is significantly associated with development of death-censored graft failure ^34^. Taken together, lower kidney functional parameters are accompanied by early timepoint tissue lipid accumulation, especially triglycerides signaling consequential structural and cellular necrosis resulting from ischemic damage. Future studies with larger cohorts are needed to validate these findings and to investigate the biological relevance to graft success.

Our study had notable strengths. The perfusate utilized in our EVNP system has been shown to be safe and has been reported in multiple settings. Extension of the perfusion period was longer than has been reported clinically. A combination of hemodynamic and functional measurements was used to determine the optimal perfusion duration using our EVNP system. We used an untargeted comprehensive metabolomics platform to identify metabolic perturbations linking poor performance and reduced kidney functional parameters during ENVP. Lastly, we assessed tissue lipid profile changes to identify potential early-timepoint lipid biomarkers that could potentially be used to predict post-transplant graft success utilizing a brief period of normothermic perfusion preservation. This study was not without limitations. Our study cohort had a relatively small sample size. Differentiation between good and poor performers was based on ex vivo hemodynamic and functional parameters which have not been validated clinically. The perfusate was based on crystalloid solution with relatively low oncotic pressure which may not be appropriate for long perfusion periods. The lack of lipid supplementation in the perfusate does not mimic the normal physiological state. Serial tissue collection used for omics assessment might have introduced tissue heterogeneity into our temporal comparisons. We are also unable to determine the impact of impaired tubular transport of amino acids on changes in metabolic profile and functional parameters. Lastly, low viability and reduced creatinine clearance among poor performers may have contributed to passive diffusion of the metabolites into the cells leading to exaggerated elevation of tissue metabolites.

In conclusion, our study showed EVNP up to 6 hours is safe, feasible and optimally distinguishes kidney function. Kidney function may be determined by metabolic flexibility characterized by BCAA metabolism. Lipid profile differences distinguishing good and poor performers occur within 1 hour with significant elevation of membrane structure components such as glycerolipids and sphingolipids in poor performers. As clinical experience with EVNP expands, particularly with application to assessment and treatment of higher risk kidneys, longer periods of perfusion should be considered to optimally assess and prepare these kidneys for transplantation. Our observation that poor performers tended to have lower KDPI, and younger age compared to good performers should prompt larger investigation on how KDPI translates into donor kidney function prior to transplantation. Further studies are necessary to better understand metabolic changes within the kidney to inform development of targeted therapeutics optimally supporting the organ during preservation and improve post-transplant outcomes.

### Disclosure

The authors of this manuscript have no conflicts of interest to disclose as described by the *Kidney International*.

## Data Availability

All data produced in the present study are available upon reasonable request to the authors

## Acknowledgments

This work was supported by a grant from the Organ Donor Research Consortium. The team would also like to thank the West Coast Metabolomics Center for their contribution to this project.

## Data sharing statement

Deidentified data, which have been stripped of all personal identification and information, will be made available to share upon request as part of the research collaboration.

---

**Supplemental Figure 1.**
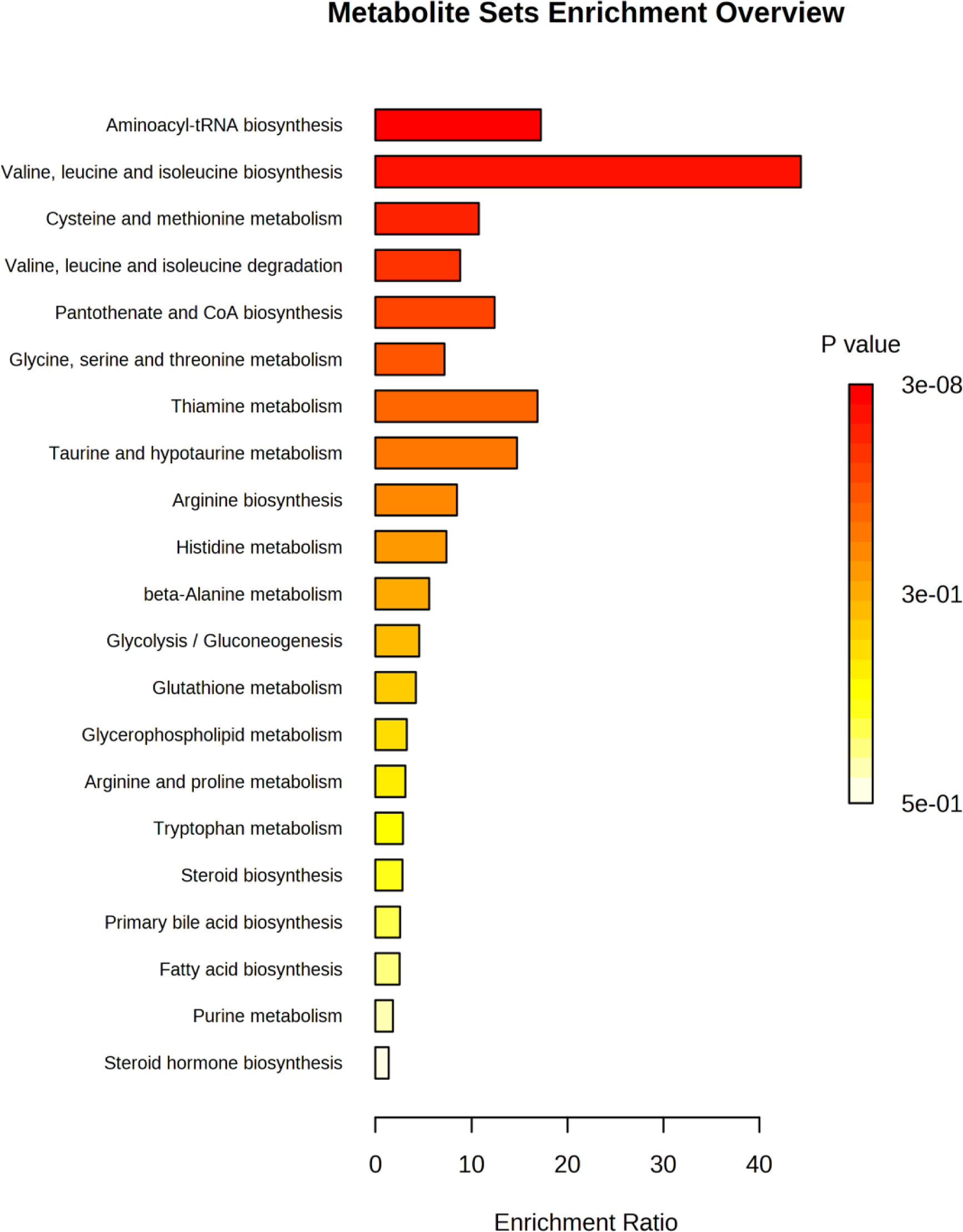
Pathway analysis of significantly altered metabolites at 6 hours during the perfusion among poor (n=4) and good performers (n=4).

**Supplemental Figure 2.**
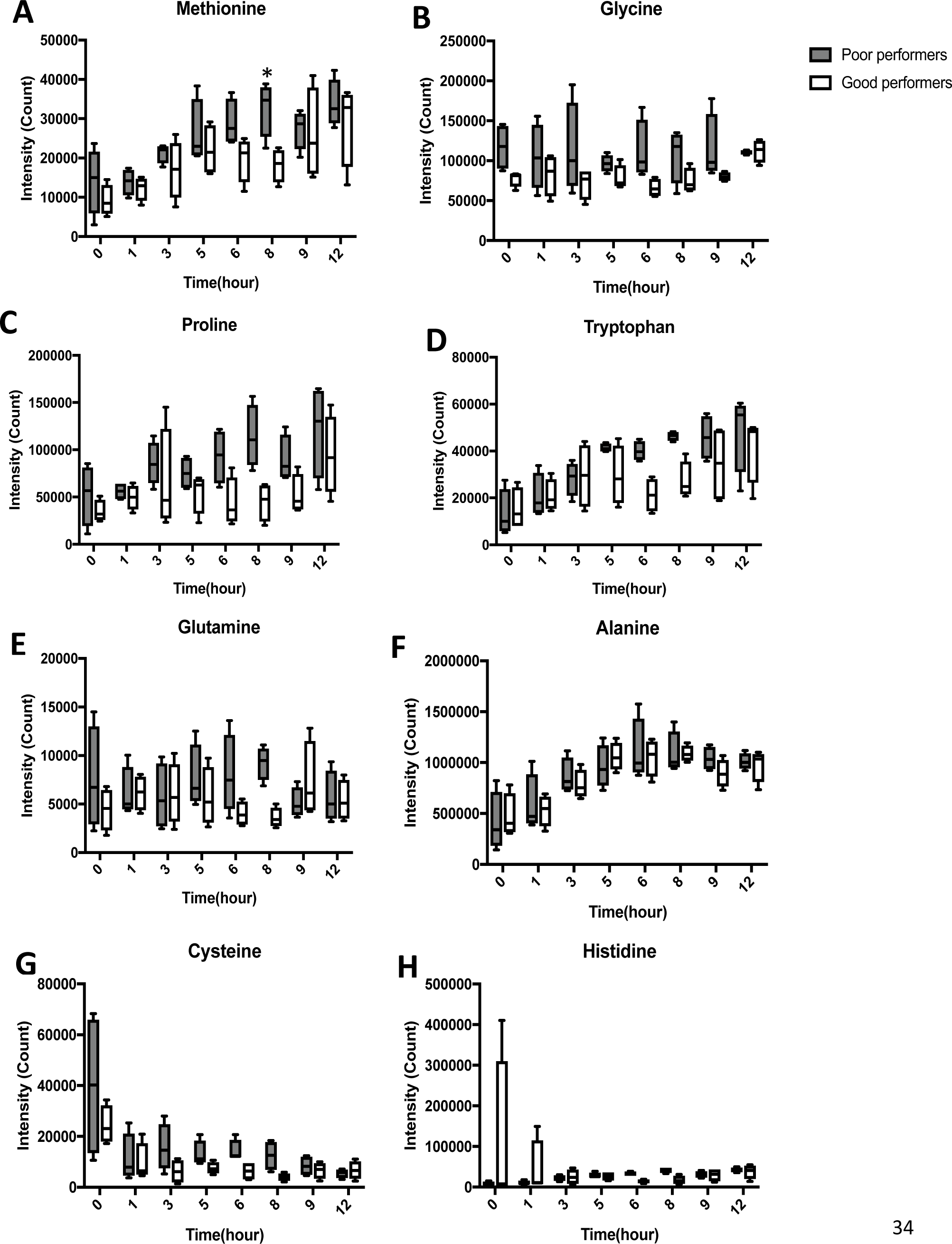
C**h**anges **in other amino acids included in the perfusion cocktail over the 12-hour perfusion period between the two groups.**

**Supplemental Figure 3.**
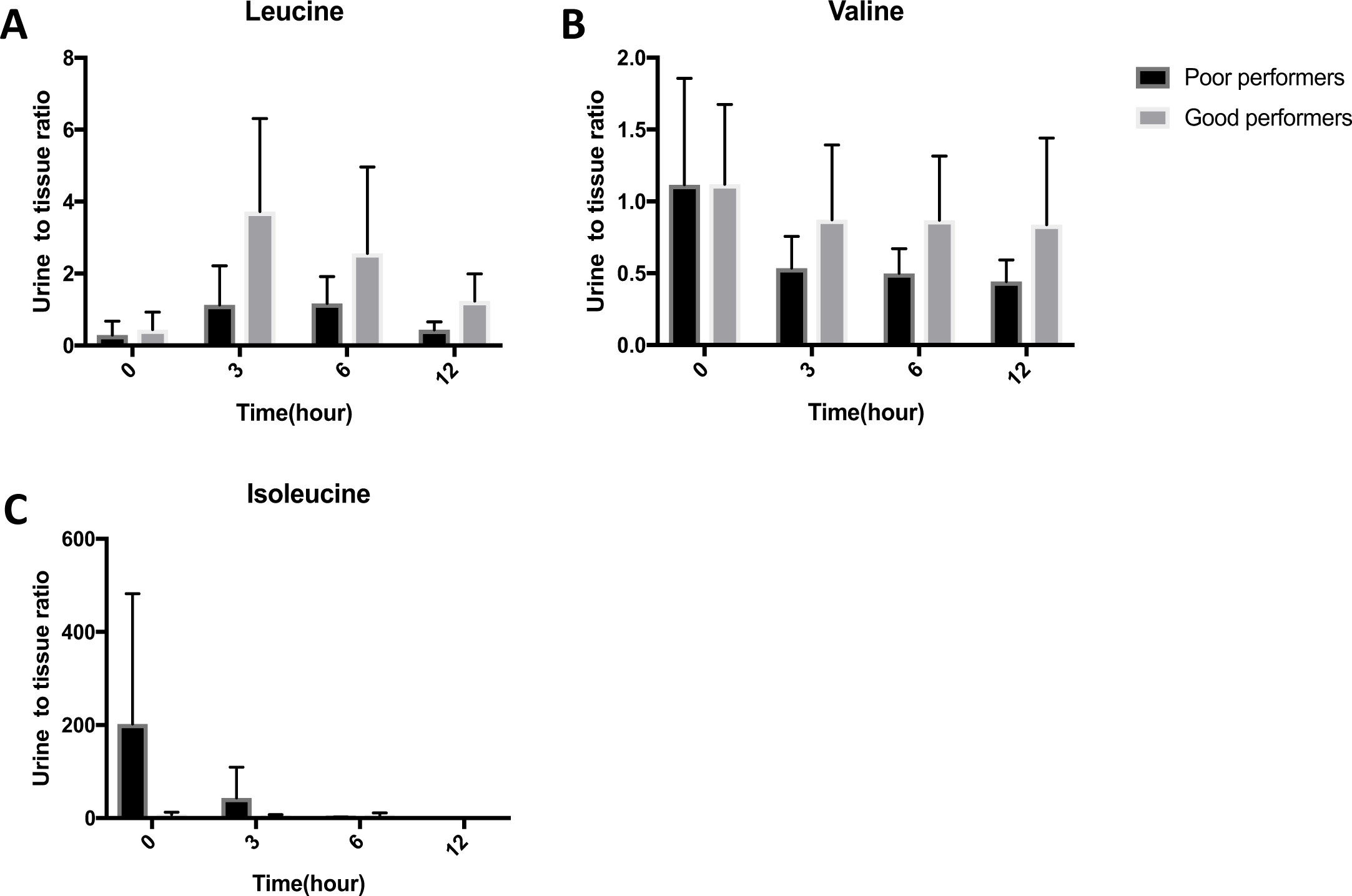
C**h**anges **in urine to tissue ratio of BCAA’s over the 12-hour perfusion period.** For panels A-C, bar graphs represent the mean and error bars represent SD.

**Supplemental Figure 4.**
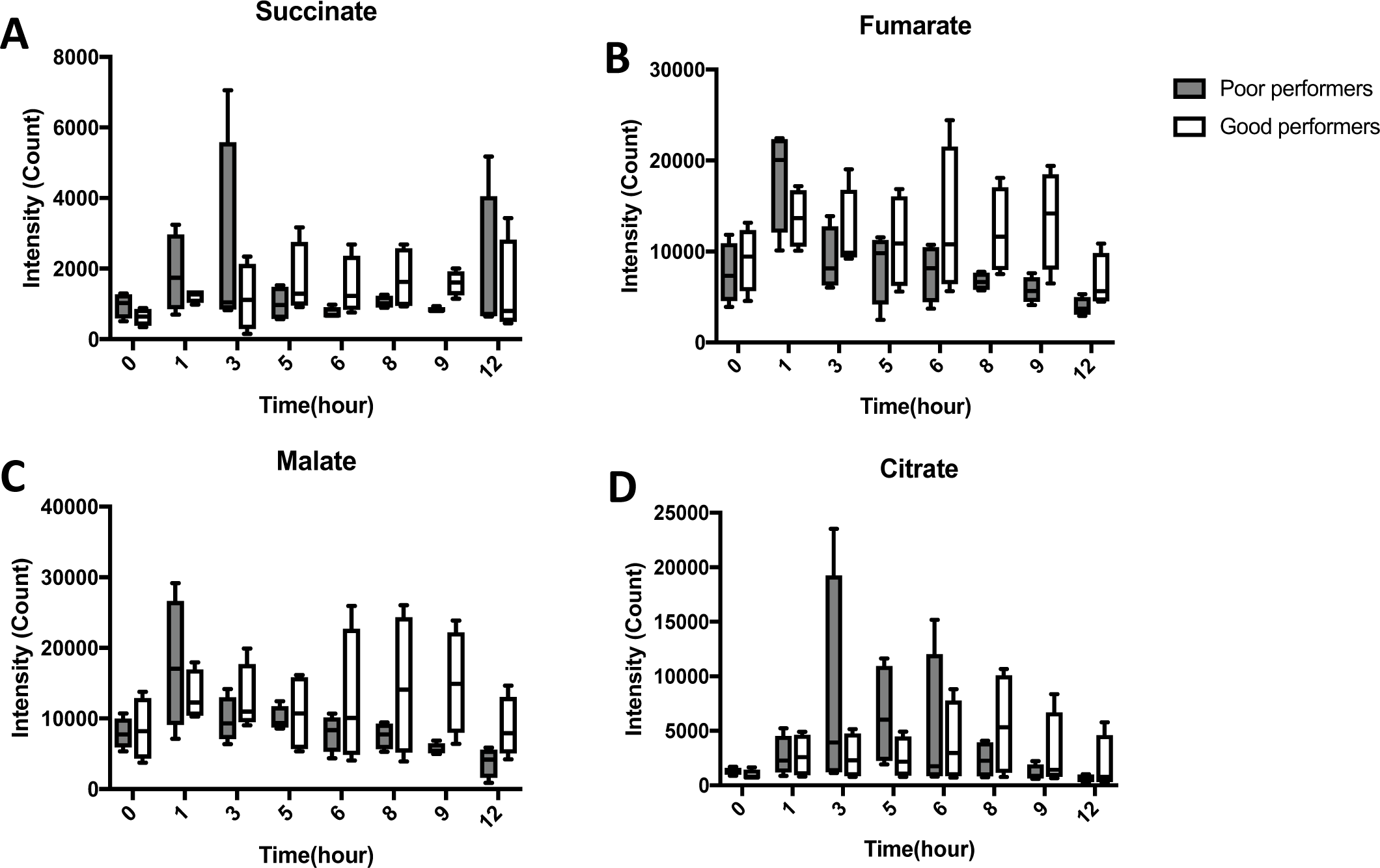
C**h**anges **in the TCA cycle intermediates during the perfusion period comparing good (n=4) and poor performers (n=4).**

**Supplemental Table 1.**
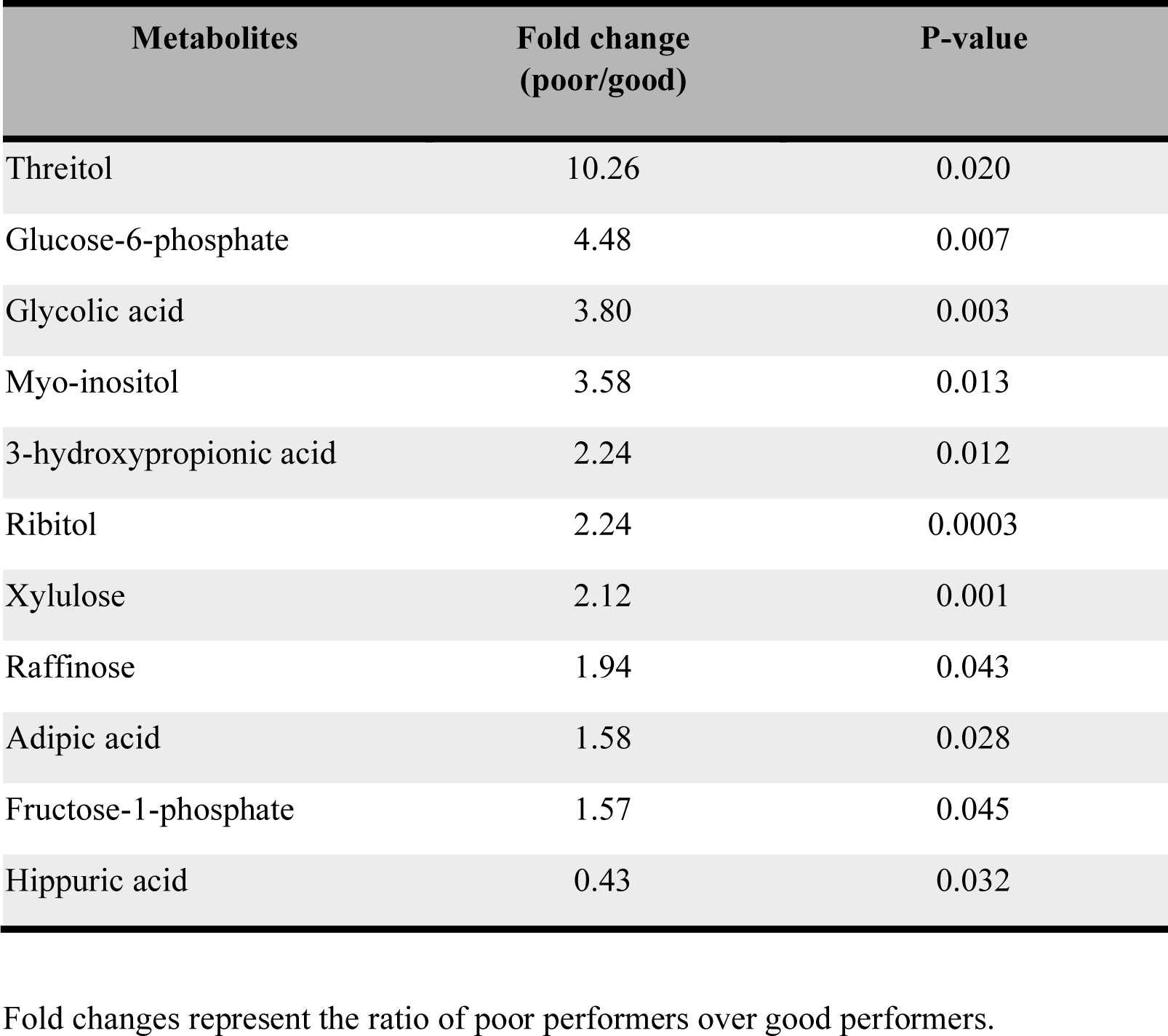
Differences in urine metabolites at 6 hours during the kidney perfusion period comparing poor (n=3) vs good performers (n=4).

**Supplemental Table 2.**
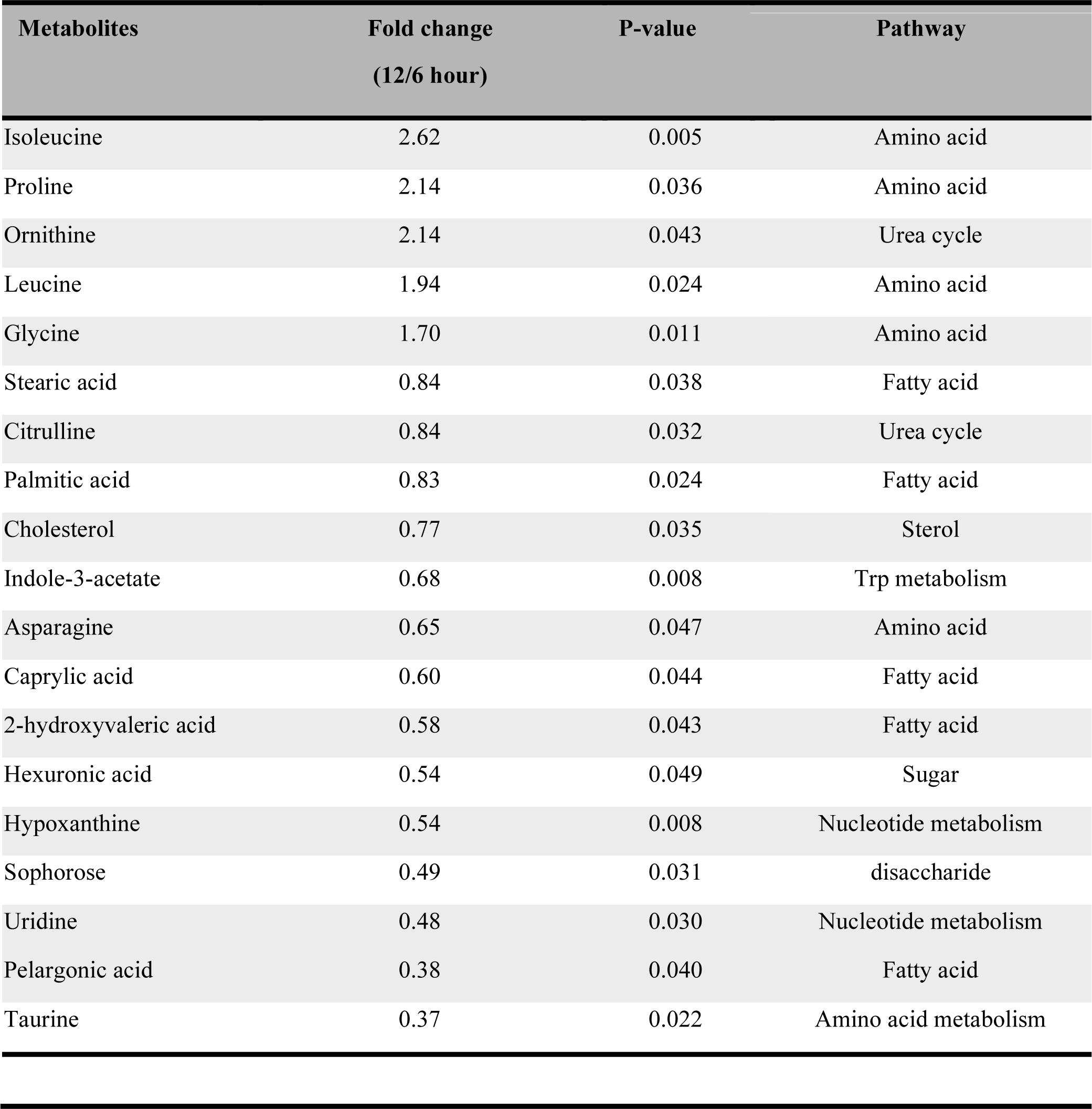
Differences in tissue metabolites among good performers comparing 12 and 6-hours post perfusion.

**Supplemental Table 3.**
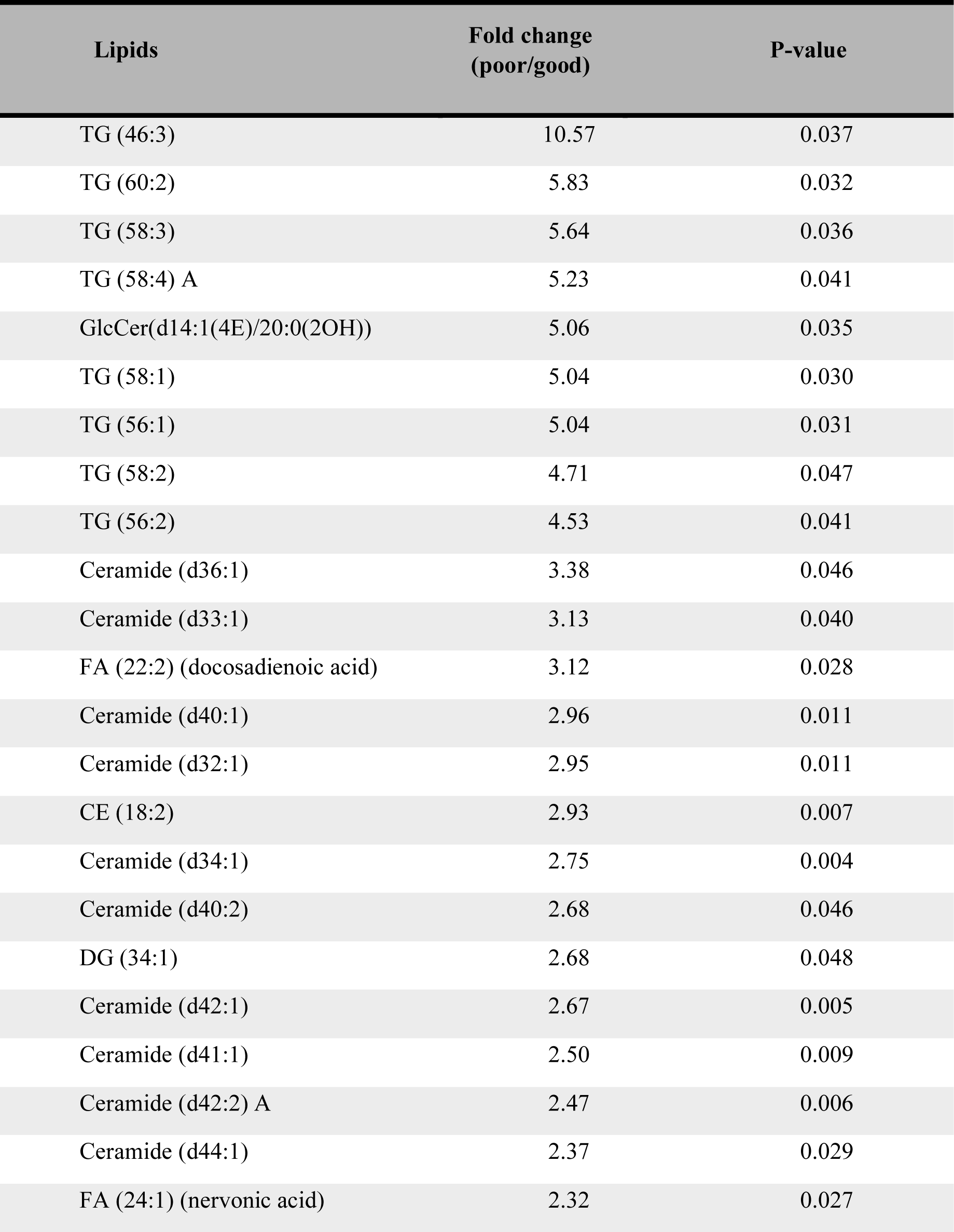
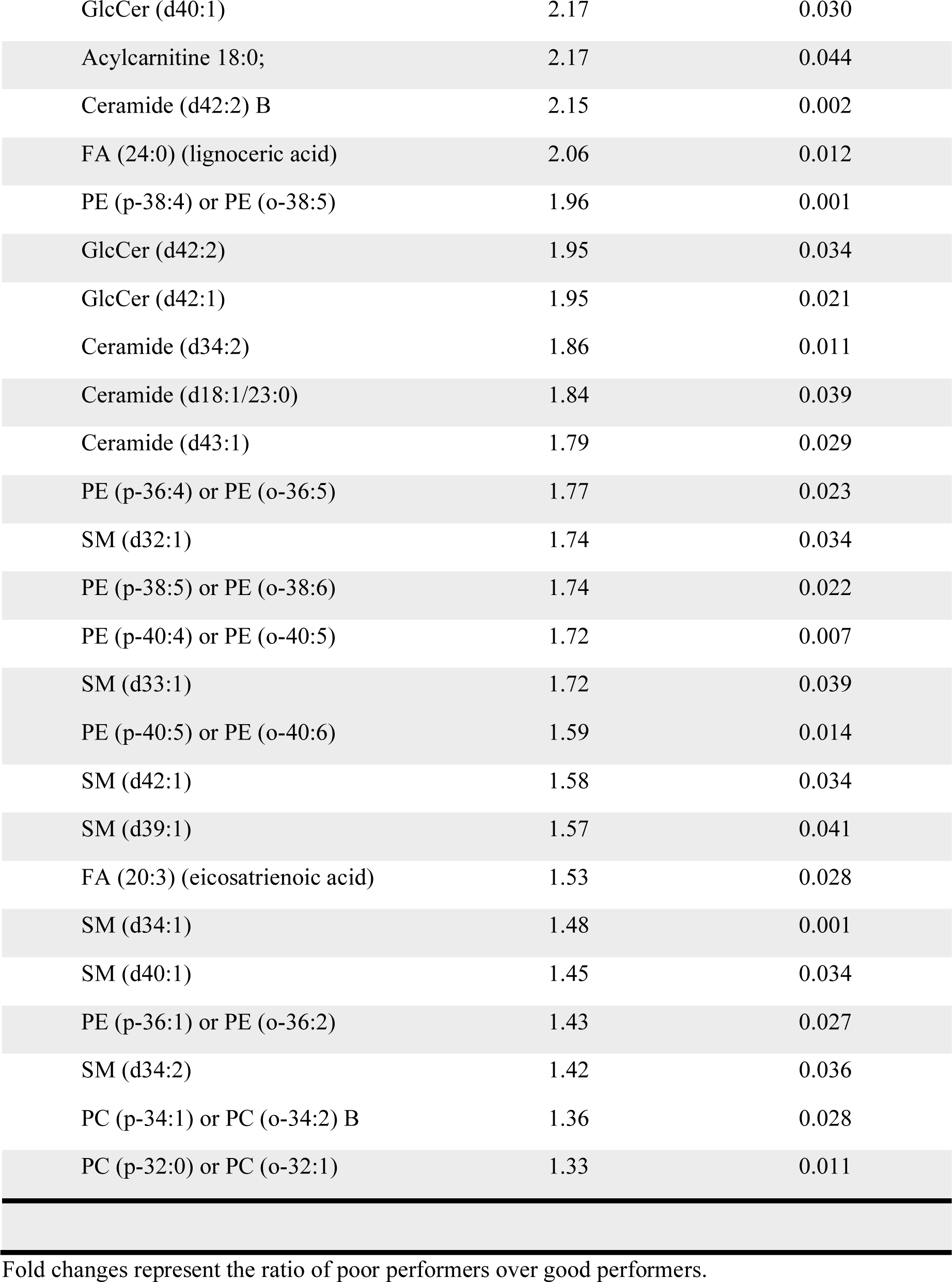
Differences in tissue lipid profile at 1-hour during the perfusion period comparing poor (n=4) vs good performers (n=4).

**Supplemental Table 4.**
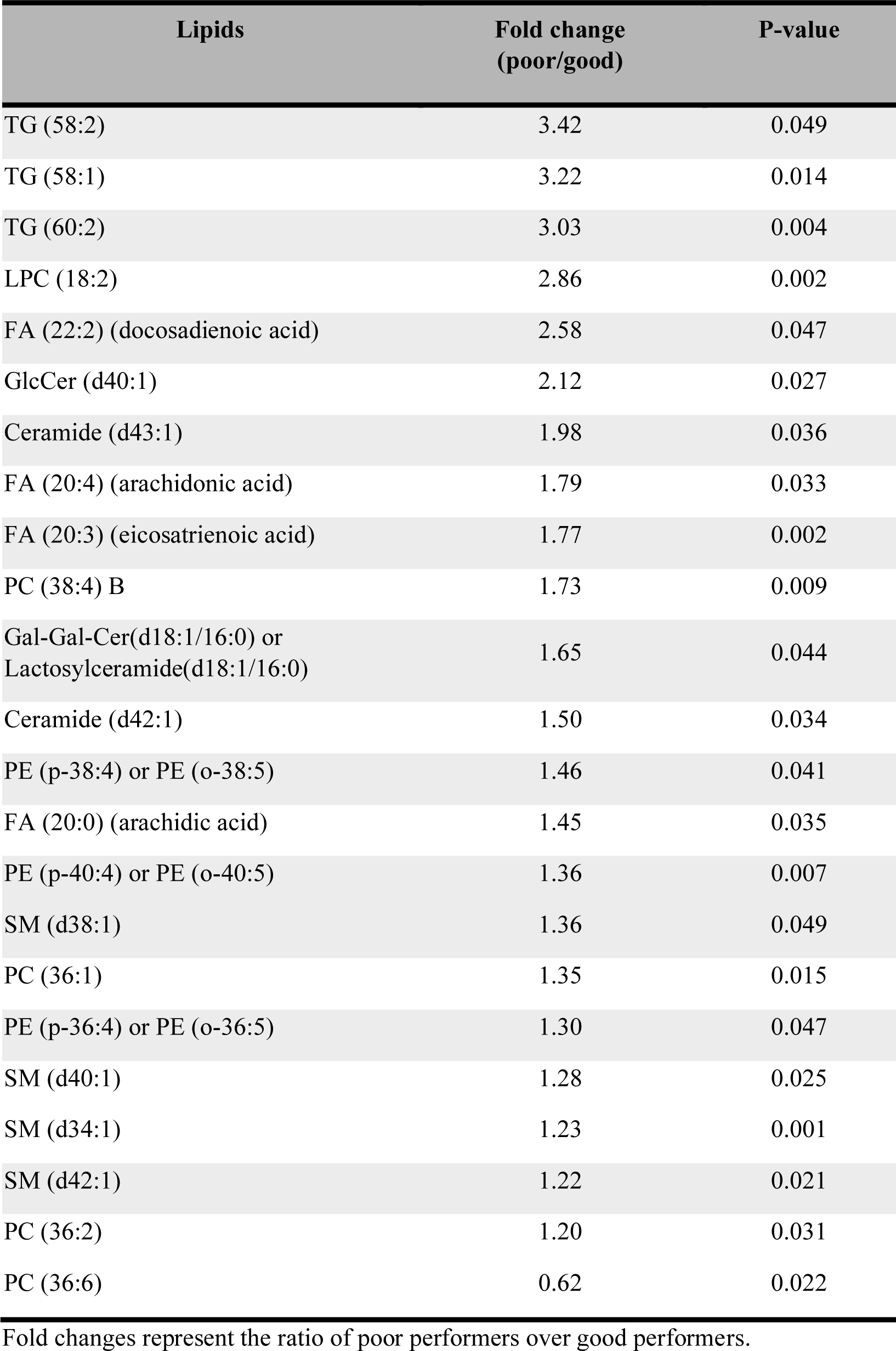
Differences in tissue lipid profile at 5-hours during the perfusion period comparing poor (n=4) vs good performers (n=4).

