## Supplemental figures for "Deceased donor kidney function is determined by branch chained amino acid metabolism during ex vivo normothermic perfusion": ENVP supplemental figures.pdf

**Supplemental Figure 1. Pathway analysis of significantly altered metabolites at 6 hours during the perfusion among poor (n=4) and good performers (n=4).**

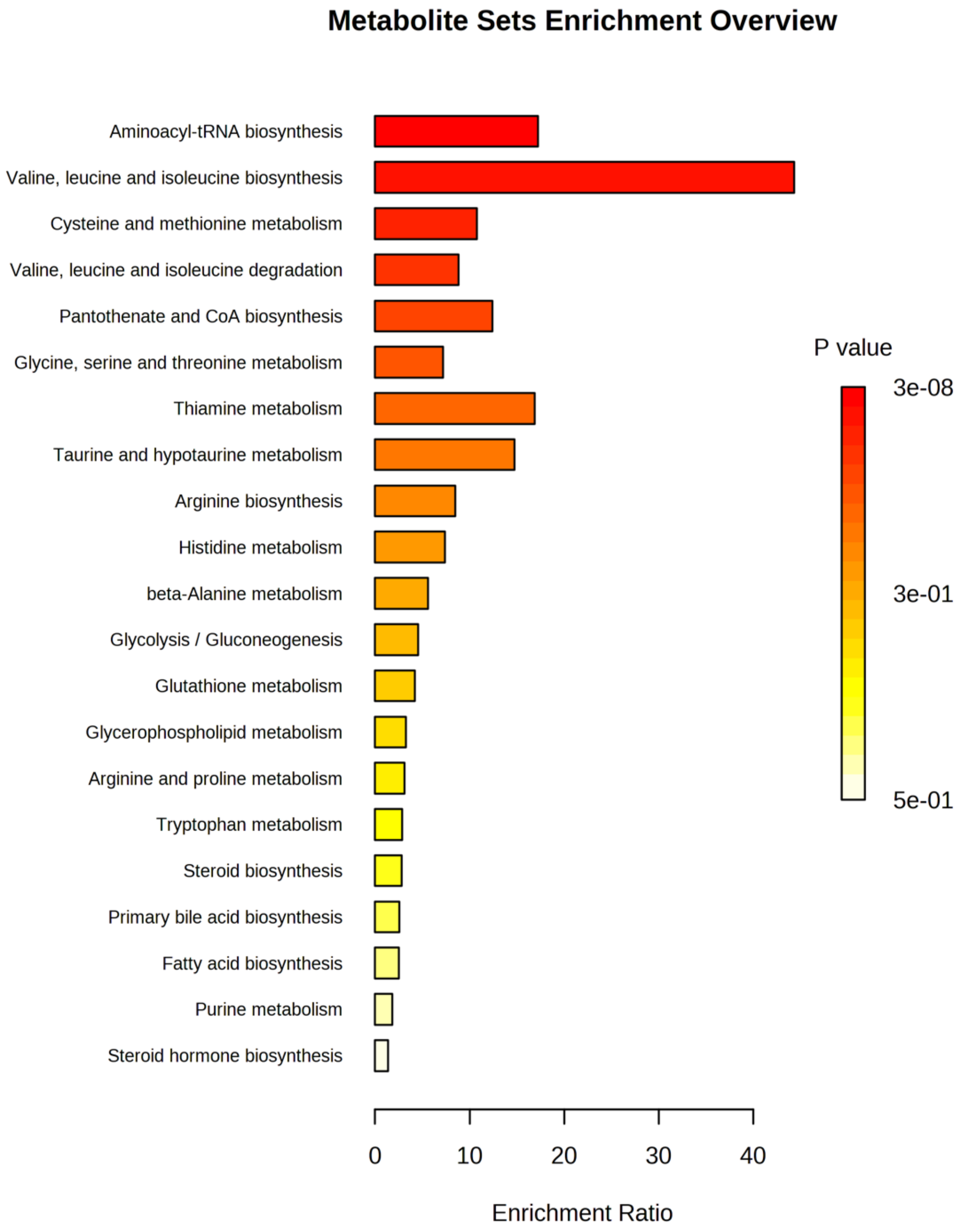

**Supplemental Table 1. Differences in urine metabolites at 6 hours during the kidney perfusion period comparing poor (n=3) vs good performers (n=4).**

| Metabolites | Fold change (poor/good) | P-value |
|---|---|---|
| Threitol | 10.26 | 0.020 |
| Glucose-6-phosphate | 4.48 | 0.007 |
| Glycolic acid | 3.80 | 0.003 |
| Myo-inositol | 3.58 | 0.013 |
| 3-hydroxypropionic acid | 2.24 | 0.012 |
| Ribitol | 2.24 | 0.0003 |
| Xylulose | 2.12 | 0.001 |
| Raffinose | 1.94 | 0.043 |
| Adipic acid | 1.58 | 0.028 |
| Fructose-1-phosphate | 1.57 | 0.045 |
| Hippuric acid | 0.43 | 0.032 |

Fold changes represent the ratio of poor performers over good performers.

**Supplemental Figure 2. Changes in other amino acids included in the perfusion cocktail over the 12-hour perfusion period between the two groups.**

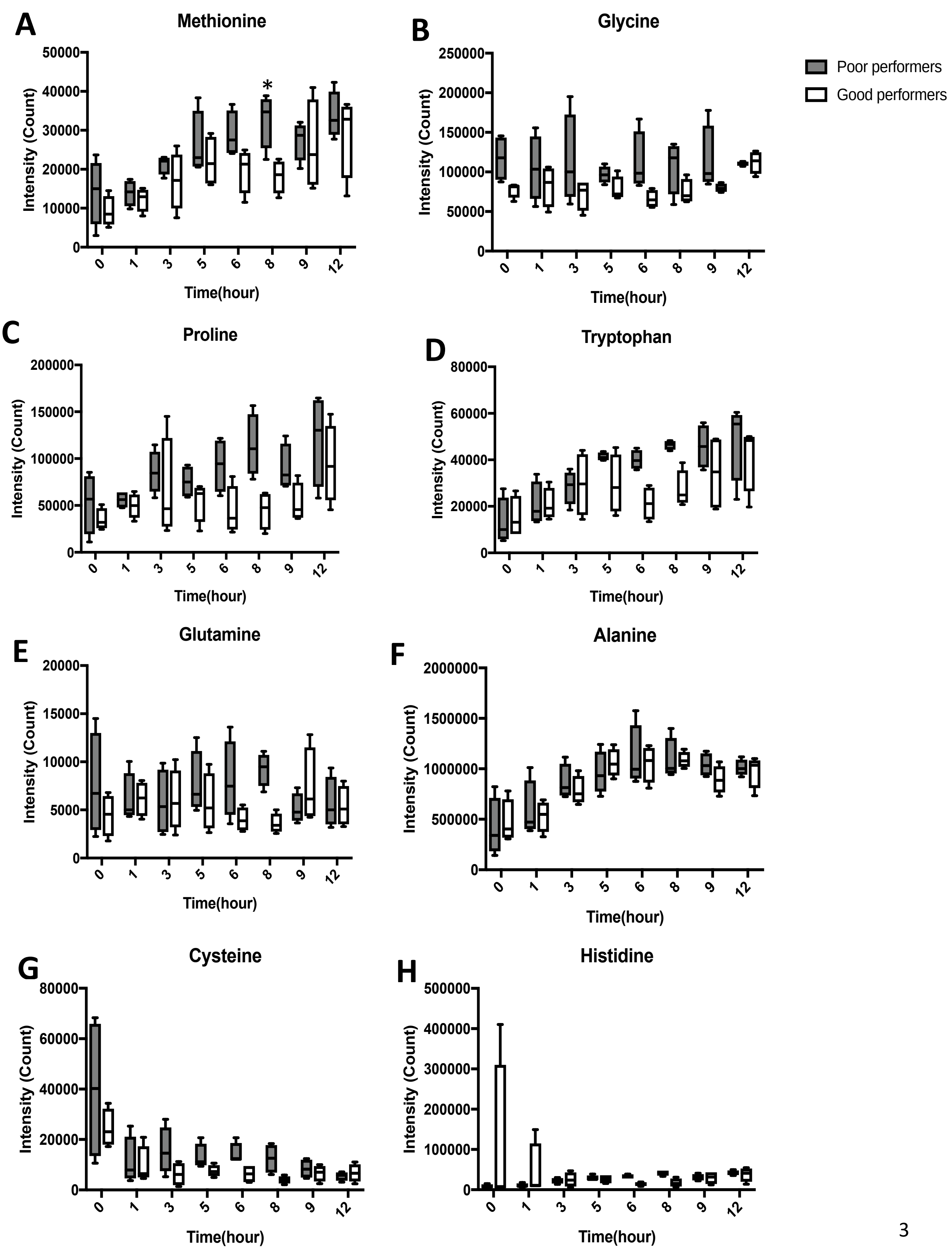

**Supplemental Figure 3. Changes in urine to tissue ratio of BCAA's over the 12-hour perfusion period.** For panels A-C, bar graphs represent the mean and error bars represent SD.

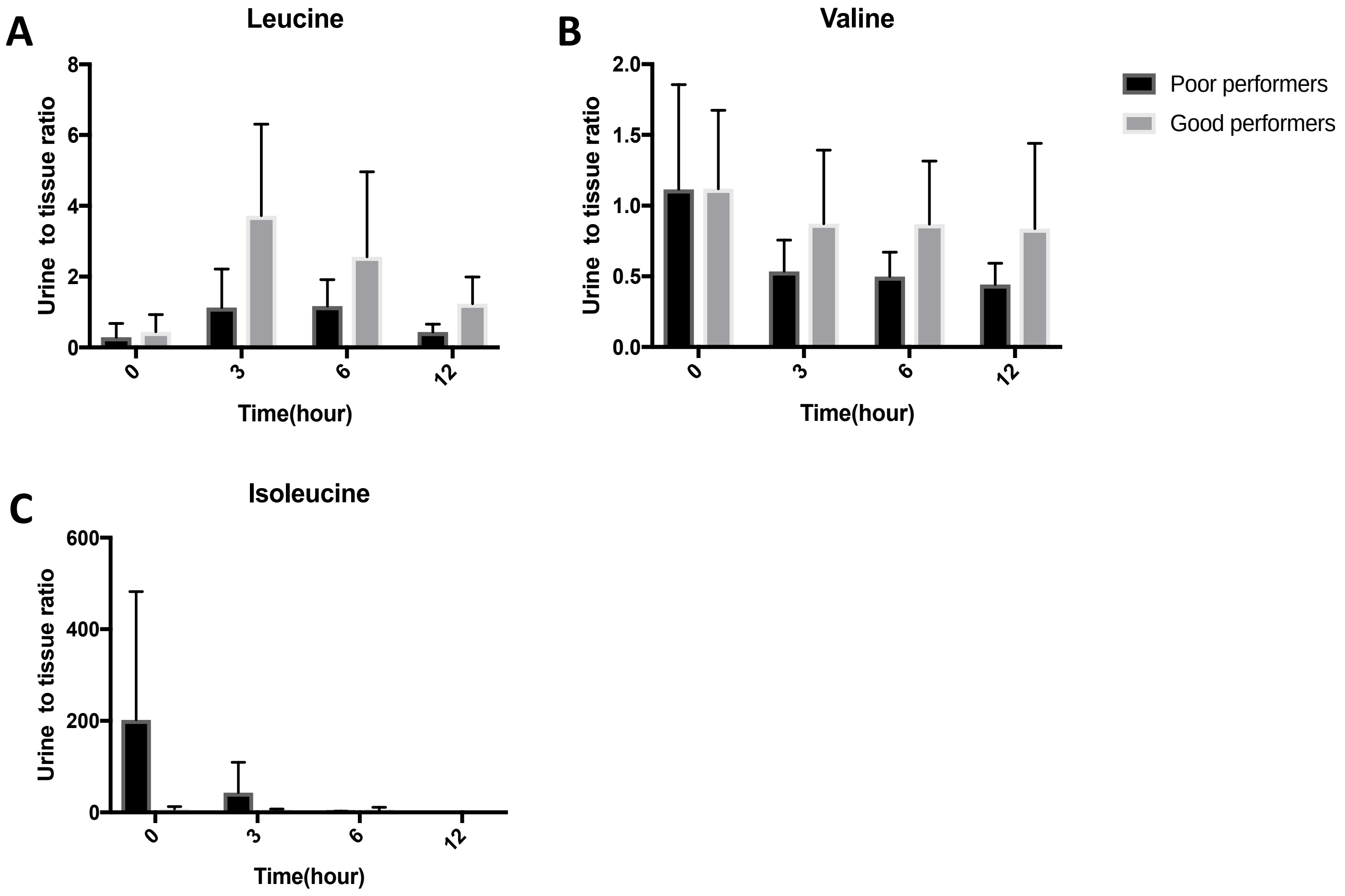

**Supplemental Table 2. Differences in tissue metabolites among good performers comparing 12 and 6-hours post perfusion.**

| Metabolites | Fold change (12/6 hour) | P-value | Pathway |
|---|---|---|---|
| Isoleucine | 2.62 | 0.005 | Amino acid |
| Proline | 2.14 | 0.036 | Amino acid |
| Ornithine | 2.14 | 0.043 | Urea cycle |
| Leucine | 1.94 | 0.024 | Amino acid |
| Glycine | 1.70 | 0.011 | Amino acid |
| Stearic acid | 0.84 | 0.038 | Fatty acid |
| Citrulline | 0.84 | 0.032 | Urea cycle |
| Palmitic acid | 0.83 | 0.024 | Fatty acid |
| Cholesterol | 0.77 | 0.035 | Sterol |
| Indole-3-acetate | 0.68 | 0.008 | Trp metabolism |
| Asparagine | 0.65 | 0.047 | Amino acid |
| Caprylic acid | 0.60 | 0.044 | Fatty acid |
| 2-hydroxyvaleric acid | 0.58 | 0.043 | Fatty acid |
| Hexuronic acid | 0.54 | 0.049 | Sugar |
| Hypoxanthine | 0.54 | 0.008 | Nucleotide metabolism |
| Sophorose | 0.49 | 0.031 | disaccharide |
| Uridine | 0.48 | 0.030 | Nucleotide metabolism |
| Pelargonic acid | 0.38 | 0.040 | Fatty acid |
| Taurine | 0.37 | 0.022 | Amino acid metabolism |

**Supplemental Figure 4. Changes in the TCA cycle intermediates during the perfusion period comparing good (n=4) and poor performers (n=4).**

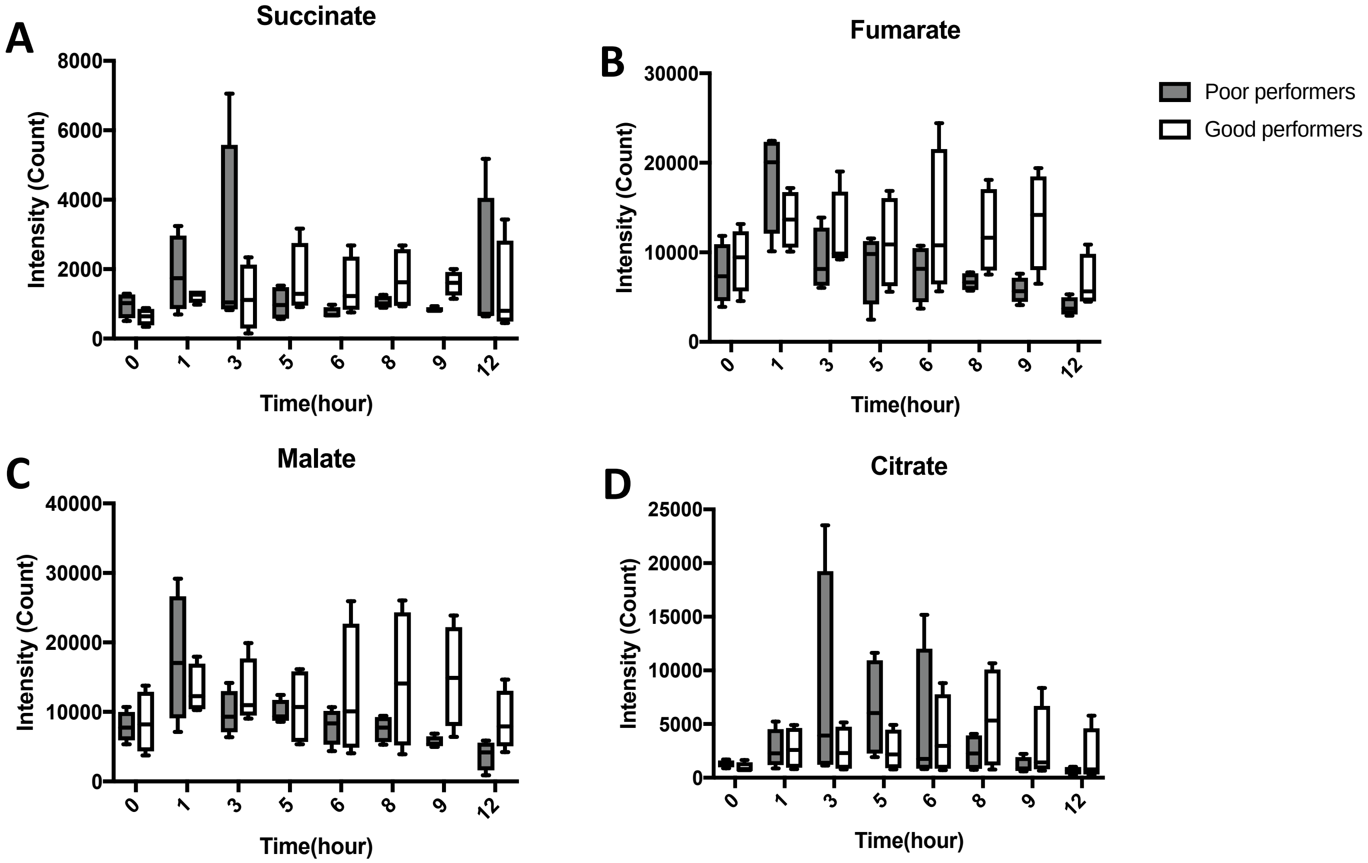

**Supplemental Table 3. Differences in tissue lipid profile at 1-hour during the perfusion period comparing poor (n=4) vs good performers (n=4).**

| Lipids | Fold change (poor/good) | P-value |
|---|---|---|
| TG (46:3) | 10.57 | 0.037 |
| TG (60:2) | 5.83 | 0.032 |
| TG (58:3) | 5.64 | 0.036 |
| TG (58:4) A | 5.23 | 0.041 |
| GlcCer(d14:1(4E)/20:0(2OH)) | 5.06 | 0.035 |
| TG (58:1) | 5.04 | 0.030 |
| TG (56:1) | 5.04 | 0.031 |
| TG (58:2) | 4.71 | 0.047 |
| TG (56:2) | 4.53 | 0.041 |
| Ceramide (d36:1) | 3.38 | 0.046 |
| Ceramide (d33:1) | 3.13 | 0.040 |
| FA (22:2) (docosadienoic acid) | 3.12 | 0.028 |
| Ceramide (d40:1) | 2.96 | 0.011 |
| Ceramide (d32:1) | 2.95 | 0.011 |
| CE (18:2) | 2.93 | 0.007 |
| Ceramide (d34:1) | 2.75 | 0.004 |
| Ceramide (d40:2) | 2.68 | 0.046 |
| DG (34:1) | 2.68 | 0.048 |
| Ceramide (d42:1) | 2.67 | 0.005 |
| Ceramide (d41:1) | 2.50 | 0.009 |
| Ceramide (d42:2) A | 2.47 | 0.006 |
| Ceramide (d44:1) | 2.37 | 0.029 |
| FA (24:1) (nervonic acid) | 2.32 | 0.027 |

| | | |
|---|---|---|
| GlcCer (d40:1) | 2.17 | 0.030 |
| Acylcarnitine 18:0; | 2.17 | 0.044 |
| Ceramide (d42:2) B | 2.15 | 0.002 |
| FA (24:0) (lignoceric acid) | 2.06 | 0.012 |
| PE (p-38:4) or PE (o-38:5) | 1.96 | 0.001 |
| GlcCer (d42:2) | 1.95 | 0.034 |
| GlcCer (d42:1) | 1.95 | 0.021 |
| Ceramide (d34:2) | 1.86 | 0.011 |
| Ceramide (d18:1/23:0) | 1.84 | 0.039 |
| Ceramide (d43:1) | 1.79 | 0.029 |
| PE (p-36:4) or PE (o-36:5) | 1.77 | 0.023 |
| SM (d32:1) | 1.74 | 0.034 |
| PE (p-38:5) or PE (o-38:6) | 1.74 | 0.022 |
| PE (p-40:4) or PE (o-40:5) | 1.72 | 0.007 |
| SM (d33:1) | 1.72 | 0.039 |
| PE (p-40:5) or PE (o-40:6) | 1.59 | 0.014 |
| SM (d42:1) | 1.58 | 0.034 |
| SM (d39:1) | 1.57 | 0.041 |
| FA (20:3) (eicosatrienoic acid) | 1.53 | 0.028 |
| SM (d34:1) | 1.48 | 0.001 |
| SM (d40:1) | 1.45 | 0.034 |
| PE (p-36:1) or PE (o-36:2) | 1.43 | 0.027 |
| SM (d34:2) | 1.42 | 0.036 |
| PC (p-34:1) or PC (o-34:2) B | 1.36 | 0.028 |
| PC (p-32:0) or PC (o-32:1) | 1.33 | 0.011 |

Fold changes represent the ratio of poor performers over good performers.

**Supplemental Table 4. Differences in tissue lipid profile at 5-hours during the perfusion period comparing poor (n=4) vs good performers (n=4).**

| Lipids | Fold change (poor/good) | P-value |
|---|---|---|
| TG (58:2) | 3.42 | 0.049 |
| TG (58:1) | 3.22 | 0.014 |
| TG (60:2) | 3.03 | 0.004 |
| LPC (18:2) | 2.86 | 0.002 |
| FA (22:2) (docosadienoic acid) | 2.58 | 0.047 |
| GlcCer (d40:1) | 2.12 | 0.027 |
| Ceramide (d43:1) | 1.98 | 0.036 |
| FA (20:4) (arachidonic acid) | 1.79 | 0.033 |
| FA (20:3) (eicosatrienoic acid) | 1.77 | 0.002 |
| PC (38:4) B | 1.73 | 0.009 |
| Gal-Gal-Cer(d18:1/16:0) or Lactosylceramide(d18:1/16:0) | 1.65 | 0.044 |
| Ceramide (d42:1) | 1.50 | 0.034 |
| PE (p-38:4) or PE (o-38:5) | 1.46 | 0.041 |
| FA (20:0) (arachidic acid) | 1.45 | 0.035 |
| PE (p-40:4) or PE (o-40:5) | 1.36 | 0.007 |
| SM (d38:1) | 1.36 | 0.049 |
| PC (36:1) | 1.35 | 0.015 |
| PE (p-36:4) or PE (o-36:5) | 1.30 | 0.047 |
| SM (d40:1) | 1.28 | 0.025 |
| SM (d34:1) | 1.23 | 0.001 |
| SM (d42:1) | 1.22 | 0.021 |
| PC (36:2) | 1.20 | 0.031 |
| PC (36:6) | 0.62 | 0.022 |

Fold changes represent the ratio of poor performers over good performers.

**Acknowledgments**

This work was supported by a grant from the Organ Donor Research Consortium. The team would also like to thank the West Coast Metabolomics Center for their contribution to this project.

### Data sharing statement

Deidentified data, which have been stripped of all personal identification and information, will be made available to share upon request as part of the research collaboration.
